# Genetic Analysis of Psychosis Biotypes: Shared Ancestry-Adjusted Polygenic Risk and Unique Genomic Associations

**DOI:** 10.1101/2024.12.05.24318404

**Authors:** Cuihua Xia, Ney Alliey-Rodriguez, Carol A. Tamminga, Matcheri S. Keshavan, Godfrey D. Pearlson, Sarah K. Keedy, Brett Clementz, Jennifer E. McDowell, David Parker, Rebekka Lencer, S. Kristian Hill, Jeffrey R. Bishop, Elena I. Ivleva, Cindy Wen, Rujia Dai, Chao Chen, Chunyu Liu, Elliot S. Gershon

**Affiliations:** MOE Key Laboratory of Rare Pediatric Diseases & Hunan Key Laboratory of Medical Genetics, School of Life Sciences, and Department of Psychiatry, The Second Xiangya Hospital, Central South University, Changsha 410000, China; Department of Psychiatry and Behavioral Neuroscience, The University of Chicago, Chicago, IL 60637, USA; Department of Human Genetics, The University of Chicago, Chicago, IL 60637, USA; Institute of Neuroscience, University of Texas Rio Grande Valley, Harlingen, TX 78550, USA; Department of Psychiatry, UT Southwestern Medical Center, Dallas, TX 75390, USA; Department of Psychiatry, Beth Israel Deaconess Medical Center and Harvard Medical School, Boston, MA 02215, USA; Departments of Psychiatry and Neuroscience, Yale University School of Medicine, New Haven, CT 06511, USA; Institute of Living, Hartford Healthcare Corp, Hartford, CT 06106, USA; Departments of Psychology and Neuroscience, BioImaging Research Center, University of Georgia, Athens, GA 30602, USA; Department of Human Genetics, Emory University School of Medicine, Atlanta, GA 30322, USA; Institute for Translational Psychiatry, Münster University, Münster 48149, Germany; Department of Psychiatry and Psychotherapy, Lübeck University, Lübeck 23538, Germany; Department of Psychology, Rosalind Franklin University of Medicine and Science, Chicago, IL 60064, USA; Department of Experimental and Clinical Pharmacology and Department of Psychiatry and Behavioral Sciences, University of Minnesota, Minneapolis, MN 55455, USA; Interdepartmental Program in Bioinformatics, University of California, Los Angeles; Los Angeles, CA 90095, USA; Department of Psychiatry, SUNY Upstate Medical University, Syracuse, NY 13210, USA; Furong Laboratory, Changsha, Hunan 410000, China; National Clinical Research Center for Mental Disorders, The Second Xiangya Hospital, Central South University, Changsha, Hunan 410000, China; National Engineering Research Center of Personalized Diagnostic and Therapeutic Technology, Central South University, Changsha, Hunan 410000, China

## Abstract

The Bipolar-Schizophrenia Network for Intermediate Phenotypes (B-SNIP) created psychosis Biotypes based on neurobiological measurements in a multi-ancestry sample. These Biotypes cut across DSM diagnoses of schizophrenia, schizoaffective disorder and bipolar disorder with psychosis. Two recently developed *post hoc* ancestry adjustment methods of Polygenic Risk Scores (PRSs) generate Ancestry-Adjusted PRSs (AAPRSs), which allow for PRS analysis of multi-ancestry samples. Applied to schizophrenia PRS, we found the Khera AAPRS method to show superior portability and comparable prediction accuracy as compared with the Ge method. The three Biotypes of psychosis disorders had similar AAPRSs across ancestries. In genomic analysis of Biotypes, 12 genes and isoforms showed significant genomic associations with specific Biotypes in Transcriptome-Wide Association Study (TWAS) of genetically regulated expression (GReX) in adult brain and fetal brain. TWAS inflation was addressed by inclusion of genotype principal components in the association analyses. Seven of these 12 genes/isoforms satisfied Mendelian Randomization (MR) criteria for putative causality, including four genes *TMEM140*, *ARTN*, *C1orf115*, *CYREN*, and three transcripts ENSG00000272941, ENSG00000257176, ENSG00000287733. These genes are enriched in the biological pathways of Rearranged during Transfection (RET) signaling, Neural Cell Adhesion Molecule 1 (NCAM1) interactions, and NCAM signaling for neurite out-growth. The specific associations with Biotypes suggest that pharmacological clinical trials and biological investigations might benefit from analyzing Biotypes separately.

## Introduction

Among the psychosis disorders, Schizophrenia (SCZ), Schizoaffective Disorder (SAD) and Bipolar Disorder (BD), there is a considerable overlap in symptoms, illness course, cognition, psychophysiology, neurobiology^1–8^, genetic susceptibility^9–12^, and transcriptome pattern^13^. The psychosis disorders show a large genetic overlap with each other and with Autism Spectrum Disorder (ASD), Attention-Deficit/Hyperactivity Disorder (ADHD), and Major Depressive Disorder (MDD), where the genetic correlation is ∼0.7 among these common psychiatric disorders^14^. Recently (2022), Bigdeli et al. found similar overlap of Polygenic Risk Scores (PRSs) for SCZ and BD, demonstrating that the PRS for each disorder predicts the other^15^, as confirmed elsewhere^16^. Historically, the first publication to introduce a PRS into genetics in 2009^17^ was on the applicability of SCZ PRS to BD.

The three psychosis Biotypes of the Bipolar-Schizophrenia Network for Intermediate Phenotypes (B-SNIP) represent a unique categorization of psychosis disorders based on shared neurobiological variation within each Biotype category, a categorization that is distinct from conventional DSM diagnostic categories^18, 19^. The Biotype categories^19^ were generated in persons with psychosis disorders by K-means clustering analysis of a series of neurobiological measurements including cognition^20–24^, eye-tracking metrics^25–28^, and electroencephalogram (EEG) measurements^29–33^ including auditory Event-Related Potentials (ERPs)^34^. The psychosis Biotypes are thus biologically distinct categories of persons with psychosis disorders, and it is hypothesized that this categorization may lead to better personalized medical care^35^.

Until now, there has not been a comparative analysis of genetic risk of illness across the Biotypes and across the several ancestries in B-SNIP participants. For genetic analysis of the three Biotypes, a Genome-Wide Association Study (GWAS) approach lacks sufficient power for the existing sample sizes^36^. PRS, however, can be applied and tested. Polygenic inheritance, a hallmark of psychosis disorders, involves the cumulative influence of numerous common Single Nucleotide Polymorphisms (SNPs) with modest effect sizes on the development of illness^37–40^. PRS summarizes individual SNP effect sizes from GWAS on the risk of illness^41, 42^, and is widely used to estimate the polygenic liability to illness at an individual level^43^.

**Non-portability of PRS among genetic ancestries** is well-known, attributable to differences in demographic relationships, allele frequencies, and local linkage disequilibrium (LD) patterns^44–50^. Various statistical calculation methods have been developed for PRS, to integrate GWAS summary statistics from diverse populations, in order to improve the prediction accuracy of case-control status in each ancestry and across ancestries^50^. PRS-CSx is a recently developed PRS method that combines GWAS discovery data from different populations, thereby leveraging the correlation of genetic effects and LD diversity across ancestries, and accounting for ancestry-specific allele frequency, LD patterns, and sample sizes in the discovery datasets. It outperforms single-population discovery methods and improves polygenic risk prediction accuracy for disease in single ancestry samples^51^. However, PRS-CSx does not generate a portable PRS for a mixed ancestry sample (see *Results* below). For personal medicine and for assessing health care risks across diverse populations, a score that is portable among ancestries would be desirable.

**Ancestry-Adjusted PRS (AAPRS)** refers to PRS that are portable across genetic ancestries, and applicable to mixed ancestry samples. Two methods for generating AAPRS, based on *post hoc* ancestry adjustment of PRS have recently become available, using genetic ancestry to calibrate PRS mean and variance^52–54^. The Khera^52^ method trains a linear regression model of PRS using genotype principal components (PCs) in the healthy control individuals within the sample as independent variables and PRS as the dependent variable. This model is then applied to the entire dataset, and the obtained residuals are the AAPRS. A related method by Ge et al.^53^ takes the 1000 Genomes (1KG) Project samples as the reference dataset to train two linear regression models on genotype PCs, and then applies the models to the target dataset to get the AAPRS. No previous study has done comparative quantitative evaluation of these methods, on their *accuracy* (Nagelkerke’s pseudo-R^2^ and area under the curve (AUC)), and *portability* (overlap among PRS density kernels of different ancestries and minimal contribution of ancestry to the AAPRSs).

**Genomic measures**: Transcriptome-Wide Association Study (TWAS) incorporates information on gene regulation from summary data on a set of markers. In comparison with SNP-based GWAS under a broad range of genetic architectures, it may enhance detection of gene associations^55^. Currently, there are emerging TWAS studies of genomic (gene-based) variation in multiple types of molecular traits, including quantitative trait loci (QTLs) for gene expression (eQTLs), isoform expression (isoQTLs), protein expression (pQTLs), histone modification (hQTL), alternative splicing (sQTL), DNA methylation (meQTL), metabolite (mQTL), and H3K27 histone acetylation (haQTLs) in multiple adult and fetal tissues^56, 57^, which are abundant resources for the prediction of a range of genetically regulated genomic events (multi-omics).

Two neurodevelopmental models of psychosis disorders have been proposed over the past thirty years^58^. First, excessive synaptic elimination or pruning in the cerebral cortex during *late* brain development, i.e. adolescence, is hypothesized as a cause of major psychoses, including SCZ^59–61^. Second, genetic studies indicate that *early* brain development, including neuronal proliferation, migration, or synapse formation is affected in SCZ. SCZ is then re-conceptualized as a neurodevelopmental disorder with psychosis as a late, potentially preventable stage of the illness^62, 63^. By leveraging the developmental brain, 2-fold improvement in colocalizations was observed for ∼60% of GWAS loci across five neuropsychiatric disorders, compared with larger adult brain functional genomic reference panels^64^. A recent review of the neurodevelopmental model of SCZ with insights from genetics, transcriptomics, and epigenomics indicates that SCZ genetic risk is dynamic and context-dependent, varying spatiotemporally throughout neurodevelopment. It might be more effective to address the early-stage perturbations in SCZ through prediction and prophylaxis in the pre-, peri-, and neonatal stages rather than during adolescence or adulthood^65^.

In this paper, we calculated SCZ PRS for B-SNIP individuals, based on multi-ancestry data, from PGC 3 SCZ (Schizophrenia and Schizoaffective Disorder) GWAS summary statistics^37^, and BD PRS based on PGC BD GWAS summary statistics. Next, the two *post hoc* ancestry adjustment methods described above were evaluated for case-control prediction performance and portability among ancestries, and the preferred adjustment method was applied (as AAPRS) to further analyses of our multi-ancestry data. We then tested AAPRS association with Biotypes.

We also imputed genomic variables from PsychENCODE eQTL and isoQTL databases of adult^66, 67^ and fetal brains^64^, and GTEx version 8 elastic net model-based sQTL results in frontal cortex^55, 68, 69^. We then tested for gene-level, isoform-level, and splicing-level expression association with Biotypes. Biological pathway enrichment and causal (mediation) analyses based on Mendelian Randomization (MR) were further performed for the significantly associated genes.

The overall study workflow is shown in Figure 1. In our results, we found the Khera AAPRS method to be preferable to the Ge method, based on superior portability and marginally equivalent accuracy. We found no significant psychosis AAPRS differences among the three Biotypes. For the multi-omics TWAS analysis in adult and fetal brains, we found twelve genes and isoforms with expression associations that differed among specific Biotypes and healthy controls, and seven of them were found to be putative causal contributors to psychosis Biotypes by MR.

**Figure 1.**
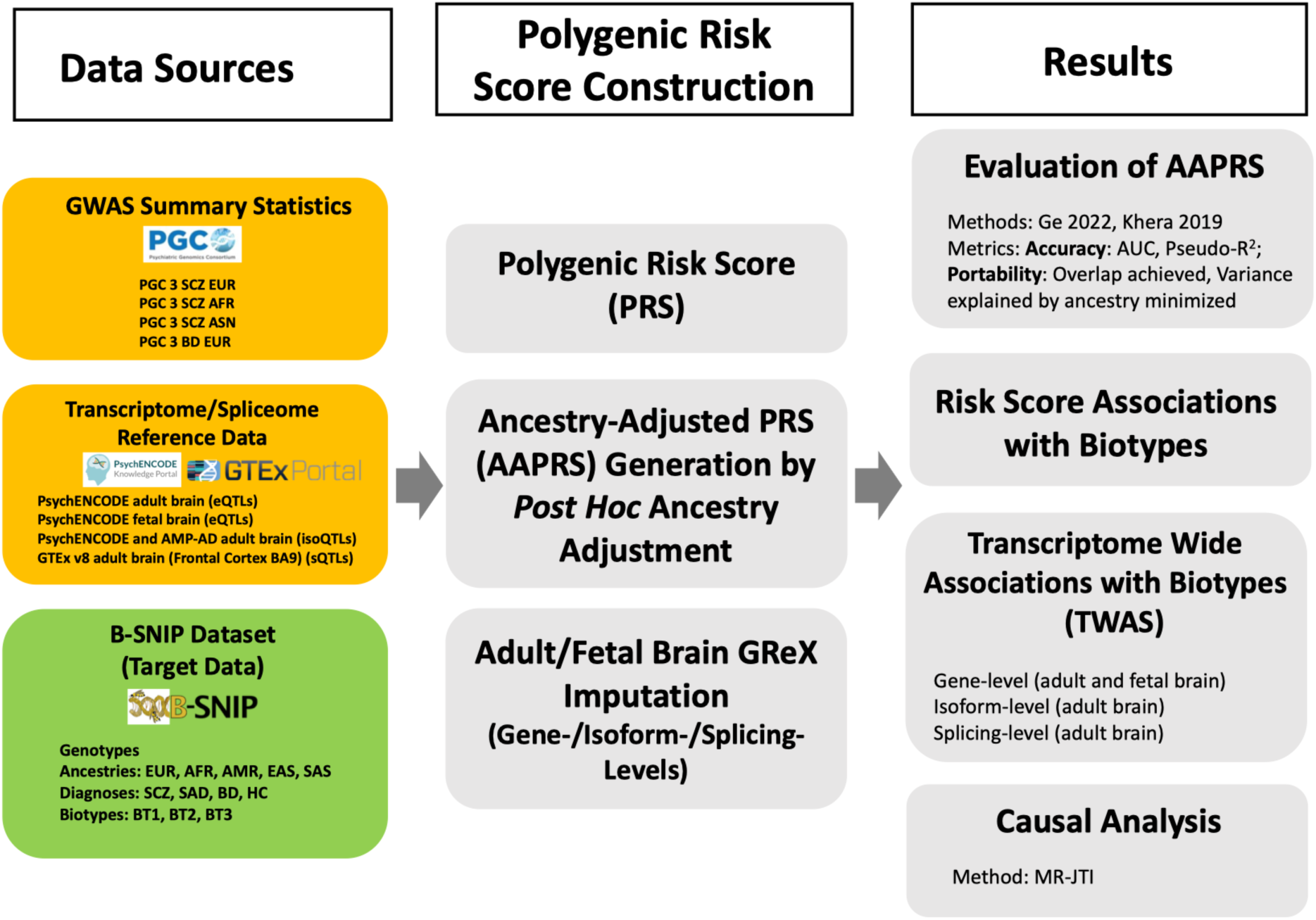
Overview of the study workflow. GWAS = Genome Wide Association Study, PRS = Polygenic Risk Score for SCZ, AAPRS = Ancestry-Adjusted Polygenic Risk Score for SCZ, TWAS = Transcriptome Wide Association Study. PGC = The Psychiatric Genomics Consortium, B-SNIP = The Bipolar and SCZ Network for Intermediate Phenotypes consortium, GTEx = The Genotype-Tissue Expression project. SCZ = Schizophrenia, SAD = Schizoaffective Disorder, BD = BD with psychotic features, HC = Healthy Control. BT1 = Biotype 1, BT2 = Biotype 2, BT3 = Biotype 3. EUR = European, AFR = African, AMR = Admixed American, EAS = East Asian, SAS = South Asian. GReX = Genetically Regulated eXpression.

## Materials and methods

### B-SNIP dataset

There are 2505 unrelated individuals who have genotypes in the B-SNIP dataset. The diagnoses are SCZ, SAD, BD, and Healthy Control (HC). Biotypes of psychosis disorders (SCZ, SAD, BD with psychosis) were obtained from our previous study^19^. Not all genotyped individuals have Biotype status or diagnosis, for reasons of incomplete phenotyping. Multiple ancestries exist in this dataset based on genotype principal component (PC) assignment of ancestry. Only previously collected data in this dataset was studied. An earlier publication documented the informed consent^70^.

### Imputation and quality control (QC) of the B-SNIP genotypes

Genotype imputation for all unrelated individuals was done using Minimac4 on the Michigan Imputation Server, taking 1KG phase 3 v5 (hg19) mixed population as the reference panel with Eagle as the phasing algorithm^71^. There are two batches (B-SNIP1 and B-SNIP2) in the B-SNIP dataset. The two batches were merged into a single dataset and QC was performed after merging.

Genetic markers were retained to have imputation quality metric R^2^ > 0.3 (which removes > 70% of poorly imputed SNPs at the cost of < 0.5% well-imputed SNPs)^72^, missingness < 0.001%, MAF > 1%, and HWE P < 1E-5. Individuals with genotype missingness > 0.05 or with Cryptic Relatedness of 2^nd^ degree or closer were filtered out using the KING program^73^. LD pruning was not done, because of the several ancestries. Genotype-based sex and heterozygosity rates were also checked for quality control. There were 10,321,126 total variants after QC.

### Ancestry assignment for the B-SNIP samples

We merged all B-SNIP unrelated samples (N = 2,505) with the 1KG phase 3 data (N = 2,504)^74^ and retained shared common variants between the two datasets. We then calculated PCs based on the LD-pruned variants (PLINK --indep-pairwise 200 100 0.1) in the merged dataset. A Random-Forest (RF) method was used, based on the first 10 genotype PCs, to assign each person in the B-SNIP dataset to one of the five 1KG super populations – European (EUR), African (AFR), Admixed American (AMR), East Asian (EAS), and South Asian (SAS).

Five ancestral groups – EUR (N = 1234), AFR (N = 908), AMR (N = 237), EAS (N = 73), and SAS (N = 53) were assigned (Figure S1). The concordance rates of RF-inferred ancestry with self-reported race in EUR, AFR, ASN (Asian: EAS + SAS), and AMR were 87%, 97%, 94%, and 16%, respectively (Table S1). The individuals (N = 2178) with both Genotype and Biotype data included 495 with Biotype 1, 483 with Biotype 2, 515 with Biotype 3, and 685 healthy controls. The detailed sample information for Biotypes on each ancestry was shown in Table S2.

### Construction of ancestry-specific PRSs

PRS-CSx was used to calculate initial (unadjusted) SCZ PRSs of each of the five ancestries (EUR, AFR, AMR, EAS, and SAS). EUR, AFR, and ASN SCZ GWAS summary statistics were used as input to generate posterior ancestry-specific SNP weights for SCZ PRSs.

PRS-CS^75^ (not PRS-CSx) was used to calculate initial (unadjusted) BD PRS due to the lack of BD GWAS summary statistics from ancestries other than EUR. EUR BD GWAS summary statistics are used as input to generate posterior EUR SNP weights for BD PRSs.

Psychosis disorder PRSs incorporating both EUR SCZ GWAS and EUR BD GWAS summary statistics were calculated using PRS-CSx *--meta* option for all EUR individuals in the B-SNIP dataset.

Ancestry-specific SCZ GWAS summary statistics were downloaded from the Psychiatric Genomics Consortium (PGC) website (https://figshare.com/articles/dataset/scz2022/19426775).

EUR BD GWAS summary statistics were downloaded from the PGC website (https://figshare.com/ndownloader/articles/14102594/versions/2). The website does not make other ancestral BD GWAS summary statistics available at this time. More detailed information about the GWAS summary statistics is shown in Table S3.

LD reference panels for each ancestry were based on 1KG Project phase 3 samples and are available at https://github.com/getian107/PRScsx.

Polymorphic variants present in all five ancestries in both B-SNIP and PGC3 SCZ GWAS datasets were used for SNP weights and PRS calculations. The parameters *--seed* and *--phi* in the PRS-CSx program were set to 1e3 and 1e-2, respectively. The posterior META SNP weights obtained from PRS-CSx program using the *--meta* option were used for PRS calculations.

Individual risk scores for each person in the B-SNIP dataset were then calculated based on the posterior META SNP weights and genotypes of B-SNIP individuals using PLINK 2.0 *--score*.

### *Post hoc* ancestry adjustment of PRS to generate Ancestry-Adjusted PRS (AAPRS)

PRS methods that can include diverse ancestries within a single dataset would increase sample size for power of detecting associations and would improve portability of PRS across ancestries. A regression-based *post hoc* ancestry adjustment method initially developed by Khera et al.^52^ was later modified by Ge et al.^53^. Both methods calculate an adjusted PRS from initial PRS as dependent variable and genotype PCs as independent variables. These linear regression models are trained on healthy control samples in the target dataset in Khera et al.^52^, and trained on 1KG data in Ge et al.^53^. The coefficients of those regressions are used in the final equations on the target dataset. These final equations normalize the PRS scores in the target dataset to generate AAPRS. These procedures can be understood as projecting the PRSs of the target dataset onto a shared space that is based on the PCs of the training dataset. For implementing the Ge AAPRS, the B-SNIP samples were projected into the 1KG phase 3 genotype PC space, and the first five genotype PCs were used for the AAPRS calculations.

To generate AAPRS for the five ancestries, we separately applied each of the two *post hoc* ancestry adjustment methods (the Khera method and the Ge method)^52, 53^ to the calculated PRS-CSx (--*meta* option) of all individuals in the B-SNIP dataset.

### Performance evaluation of the *post hoc* ancestry adjustment methods for accuracy and portability

Prediction accuracy for case-control status was evaluated by 1) the Area Under the Curve (AUC) of the Receiver Operating Characteristic (ROC), and 2) Nagelkerke’s pseudo-R-squared^76, 77^. Portability metrics were 1) Overlap of the AAPRS density kernels across ancestries, and 2) Percentage of AAPRS variance attributable to ancestry.

An implicit assumption of the *post hoc* ancestry adjustment methods is that ancestry variation is a “nuisance variable” so that a complete overlap of ancestries would generate a valid portable PRS for each person. The overlapping area among the kernel density estimates of AAPRS for the five ancestries was calculated by the *boot.overlap()* function in the ‘overlapping’ package (version 2.1) in R 4.2.1^78, 79^.

The percentage of PRS variance attributable to ancestry and to Biotype, with and without ancestry adjustment, was assessed by two-way Analysis of Variance (ANOVA) with interaction effect estimated, using the *aov()* function in R 4.2.1.

An “overfitting” question applies to the Khera method^52^, where the healthy control individuals in the target dataset are used to train the linear regression model in the first step of the adjustment. This might generate overfitting when applying the model to the entire target dataset. To test for overfitting, two-fold cross-validation was used to recompute the AAPRS and overlap statistics between the five ancestries. Specifically, we began by randomly dividing the B-SNIP dataset into two equal splits: split 1 serves as the training set and split 2 as the test set. We then reversed the roles of the two splits in a second analysis. To calculate the Khera AAPRS, we trained the Khera model on the training set and then applied it to the test set. The calculated Khera AAPRSs in two splits were then combined to calculate the overlap statistic. We compared this overlap statistic with the initial overlap statistic of Khera AAPRS to determine whether the overfitting exists.

### Comparison of PRSs and AAPRSs among Biotypes

To have uniform scales for PRSs across ancestries, percentile transformation was performed on the PRSs and AAPRSs. Wilcoxon tests were then performed on the percentile transformed PRSs or AAPRSs to assess whether there are significant PRS differences between healthy controls and Biotypes, as well as among the three different Biotypes. Bonferroni correction for six tests (N = 6) was set for statistical significance (P < 8.33e-03).

### Gene expression, mRNA isoform and splicing TWASs in adult and fetal brains

TWAS was performed for Biotypes both within and across ancestries. The PsychENCODE multi-ancestry-based adult brain^66, 67^ and fetal brain^64^ eQTL model results were used for gene-level TWAS. The isoform prediction database for isoform-level TWAS was obtained from Arjun Bhattacharya, et al.^80^. The data used to construct this prediction model were adult brain cortex tissue from 2,365 individuals compiled and processed from the PsychENCODE Consortium^66^ and the Accelerating Medicines Partnership Program for Alzheimer’s Disease (AMP-AD)^81^. GTEx version 8 elastic net model-based sQTL results in frontal cortex^55, 68, 69^ in adult brains were used for splicing-level TWAS. Detailed information about the PredictDB databases used in this study was shown in Table S4.

Using MetaXcan^55, 68^, we first imputed transcriptome, isoform and spliceome data for B-SNIP individuals. Next, the genetically regulated expression (GReX) of 14,188 genes in adult brain, 7,024 genes in fetal brain, 34,169 isoforms in adult brain, and 7,425 splicing events in adult brain were imputed for each person. Each gene/isoform/splicing in adult or fetal brain imputed GReX component was then tested for association with case (including all the three Biotypes) vs. control, SCZ (including SCZ and SAD) vs. control, BD vs. control, SCZ vs. SAD, SCZ vs. BD, SAD vs. BD, Biotype vs. control, Biotype vs. Biotype, and within and across ancestries, using a logistic regression model with the first five genotype PCs as covariates. Multiple testing significance thresholds for TWAS associations were Benjamini & Hochberg (BH)^82^ FDR correction (FDR < 0.05).

To consider possible inflation in the different TWAS models^83–85^, we used two methods to evaluate our TWAS results: 1) Genomic inflation factor *lambda*, and 2) Bayesian method-based estimated inflation using the empirical null distribution^83^. The calculations were implemented by ‘QCEWAS’ and ‘bacon’ packages in R 4.2.1. The first five genotype PCs of B-SNIP individuals were used as covariates in our main TWAS association analysis. Inflation was tested for the association analysis results under the different TWAS models with and without genotype PCs as covariates. Quantile-Quantile (Q-Q) plots and the two inflation statistics were used for visualizing and analyzing the inflation based on the observed P values in TWAS results.

### Causal analysis of candidate risk genes for psychosis Biotypes and psychosis disorders

We applied the Mendelian Randomization - Joint Tissue Imputation (MR-JTI)^86^ approach to test for putatively causal genes of psychosis Biotypes and diagnoses from TWAS results. Although MR analysis of disease is a test of causality, it is calculated as a test of the relative strength of direct and indirect associations between an event (G in Figure S2) and a [disease] outcome (D in Figure S2). The alternative to a direct association is an indirect association of G on disease outcome D, detected by a stronger association of G to another event (such as an IP) that itself is strongly associated with D. That is, the effect of G on D is indirect when the G association with the other event is stronger than the association of G with the disease (D). The relative strengths of the associations are measured as in the diagram. Bonferroni corrections are used for multiple testing of genes.

When concluding there is a causal association between events G and D, there is no conclusion on the relative strength of this causal event vs. other causal events that are not measured or reported, such as trauma or societal stresses. Nor is there a biological mechanism proven by the association, although one might be implied by the nature of the genetic association.

### Biological pathway enrichment analysis

We did biological pathway enrichment analysis for candidate genes using Reactome^87^ (https://reactome.org/).

### Resampling for robustness

Resampling was used to test the robustness of the findings from PRS and TWAS analyses of psychosis Biotypes, due to the lack of external psychosis Biotype data. Specifically, we randomly selected 80% of the whole sample for 10 times, and each time we performed all the PRS and TWAS analyses. We then evaluated the robustness of our findings by comparing the results from resampling and from our main analyses.

## Results

### 1. Polygenic Risk Scores

#### Similar prediction accuracy for case-control status from both AAPRS methods

In the combined multi-ancestry sample, the unadjusted PRS gave 0.606 (95% CI: 0.580, 0.631) for AUC and 0.043 (95% CI: 0.021, 0.062) for Nagelkerke’s pseudo-R^2^ on case-control status. The Ge method gave 0.619 (95% CI: 0.594, 0.644) for AUC and 0.053 (95% CI: 0.028, 0.075) for Nagelkerke’s pseudo-R^2^, and the Khera method gave 0.607 (95% CI: 0.582, 0.632) and 0.044 (95% CI: 0.021, 0.063) respectively. Thus, these two AAPRS methods performed comparably to the unadjusted PRS on both AUC and Nagelkerke’s pseudo-R^2^ for case-control status in the combined multi-ancestry sample and in each of the five ancestries separately (Figure 2, Table S5).

**Figure 2.**
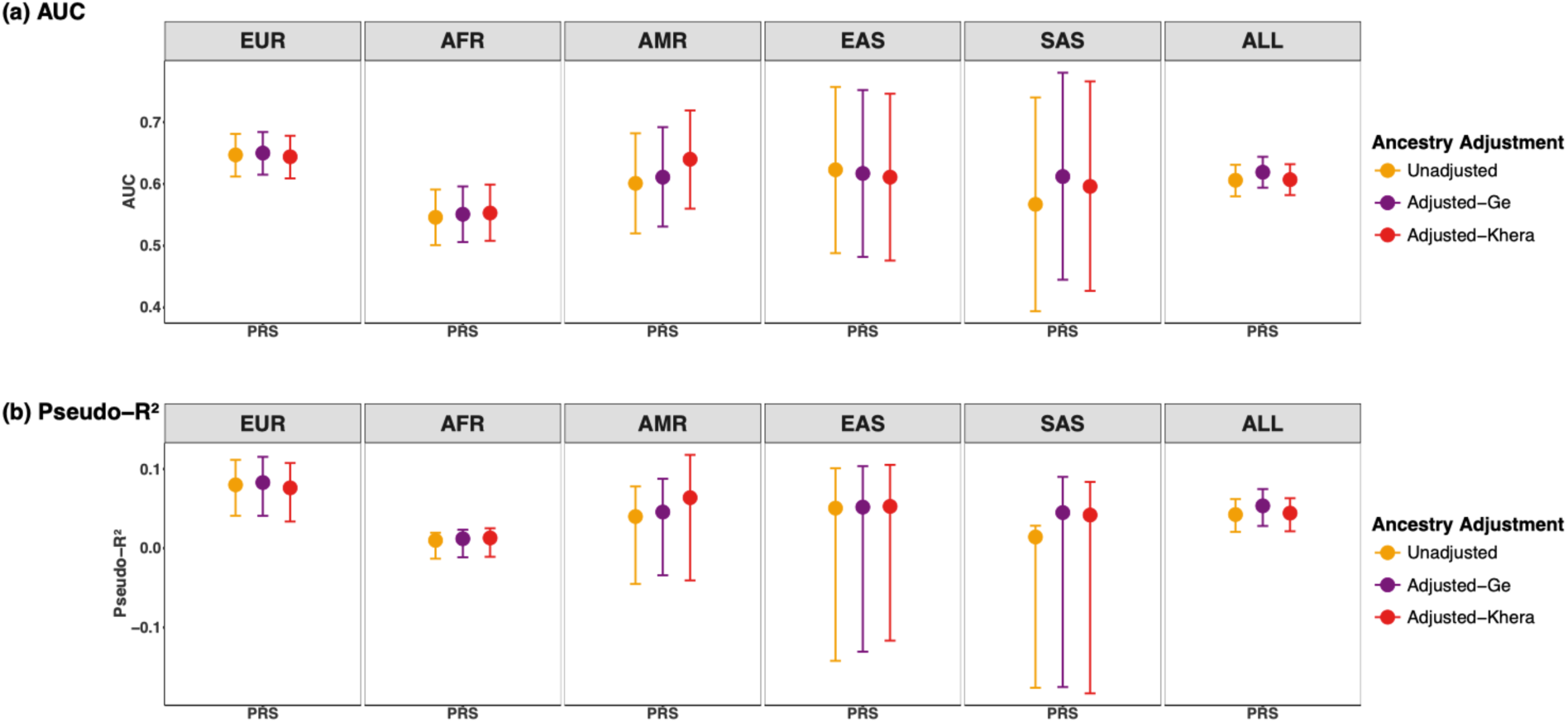
Prediction accuracy of PRSs for case-control status before and after ancestry adjustment within and across 5 ancestries. All the PRSs are calculated based on EUR, AFR and Asian SCZ GWAS summary statistics. (a) The area under the receiver operating characteristic (ROC) curve (AUC) of PRSs. (b) The proportion of the case-control variance (Nagelkerke’s pseudo-R^2^) explained by PRSs. Lines for Nagelkerke’s pseudo-R^2^ in (b) correspond to 95% confidence intervals calculated via 1000 bootstrapping. The five ancestries were assigned by Random Forest inferred method based on 1KG reference. EUR = European, AFR = African, AMR = Admixed American, EAS = East Asian, SAS = South Asian, ALL = Combined multi-ancestry individuals of all the five ancestries. “Unadjusted” risk scores are the *--meta* option results from PRS-CSx prior to *post hoc* ancestry adjustment, and “Adjusted” refers to AAPRS (Ancestry-Adjusted Polygenic Risk Score) with *post hoc* ancestry adjustment of Khera or Ge. We find no overall advantage in prediction accuracy of case-control status for either adjustment method.

#### On PRS overlap across ancestries, the Khera AAPRS gives greater improvement

Overlap of PRS density kernels across different ancestries is one of the portability metrics for the AAPRS evaluation. The unstated assumption in reducing overlap is that there are no true differences between ancestry PRSs, and they should overlap completely.

The estimated overlap of unadjusted PRSs among the five ancestries was 0.914 (95% CI: 0.902, 0.926). The estimated overlap of the Ge AAPRS and the Khera AAPRS was 0.962 (95% CI: 0.955, 0.969) and 0.974 (95% CI: 0.966, 0.982) respectively. Both AAPRS methods improved overlap compared to that of unadjusted PRS, and the improvement was greater with the Khera adjustment than with the Ge adjustment, while complete overlap was not observed in our data (Figure 3).

**Figure 3.**
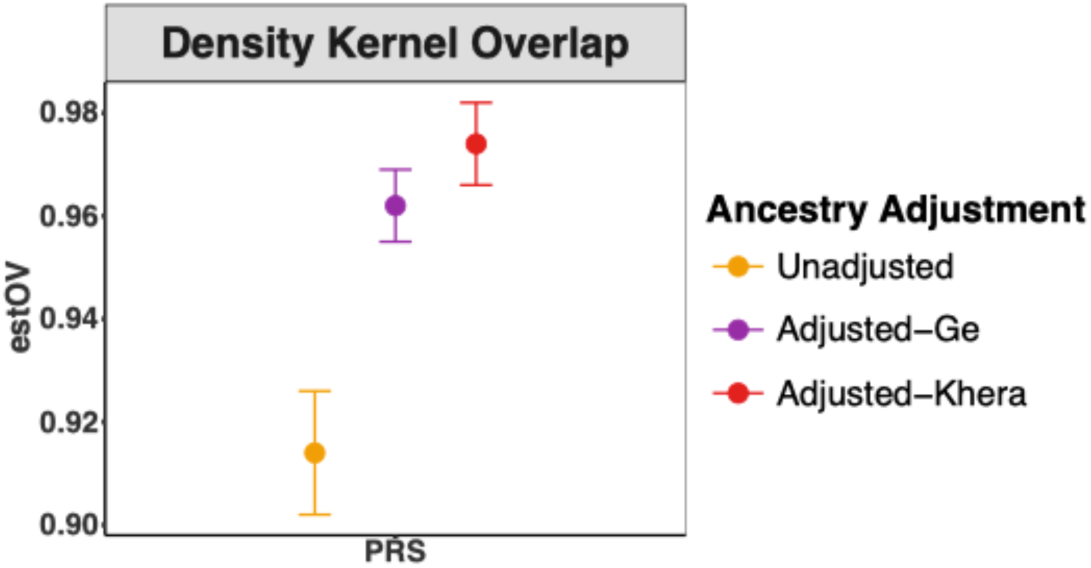
Effects of *post hoc* ancestry adjustment on the overlap of PRS-CSx (meta option) distributions among 5 ancestries. Figure shows the overlaps of density kernels of PRSs between different ancestries before and after ancestry adjustment. ‘estOV’ = estimated overlapping area of risk scores between different ancestries. Standard error of estOV is calculated by 1000 bootstrap draws (meaning 1000 iterations with bootstrapping), and the labelled error bar of upper and lower values is estOV +/- SE. Between the two PRS *post hoc* ancestry adjustment methods, Khera adjustment gave greater PRS overlap (97% vs. 96%) between different ancestries, both significantly higher than the overlap (91%) of unadjusted PRSs.

#### The Khera method shows smaller ancestry contribution than the Ge method to the variance of AAPRS

Two-way analyses of variance (ANOVA) of the combined sample were performed to assess PRS variance contributions of Biotype, Ancestry, and residuals. As expected, the method that yielded the greatest PRS density kernel overlap among five ancestries (Khera) showed the smallest contribution of ancestry to AAPRS (Figure 4, Table S6). Even there, we found ancestry contributions to AAPRS. With the Khera method, ancestry accounted for 1% (P = 4.37e-04) of AAPRS variance versus 14% (P = 3.03e-70) for Ge. There were no significant interactions between Biotype and ancestry in either Ge AAPRS or Khera AAPRS (Figure 4, Table S6). The Khera result thus more closely fits the unstated assumption of ancestry as a nuisance variable (this assumption is discussed below).

**Figure 4.**
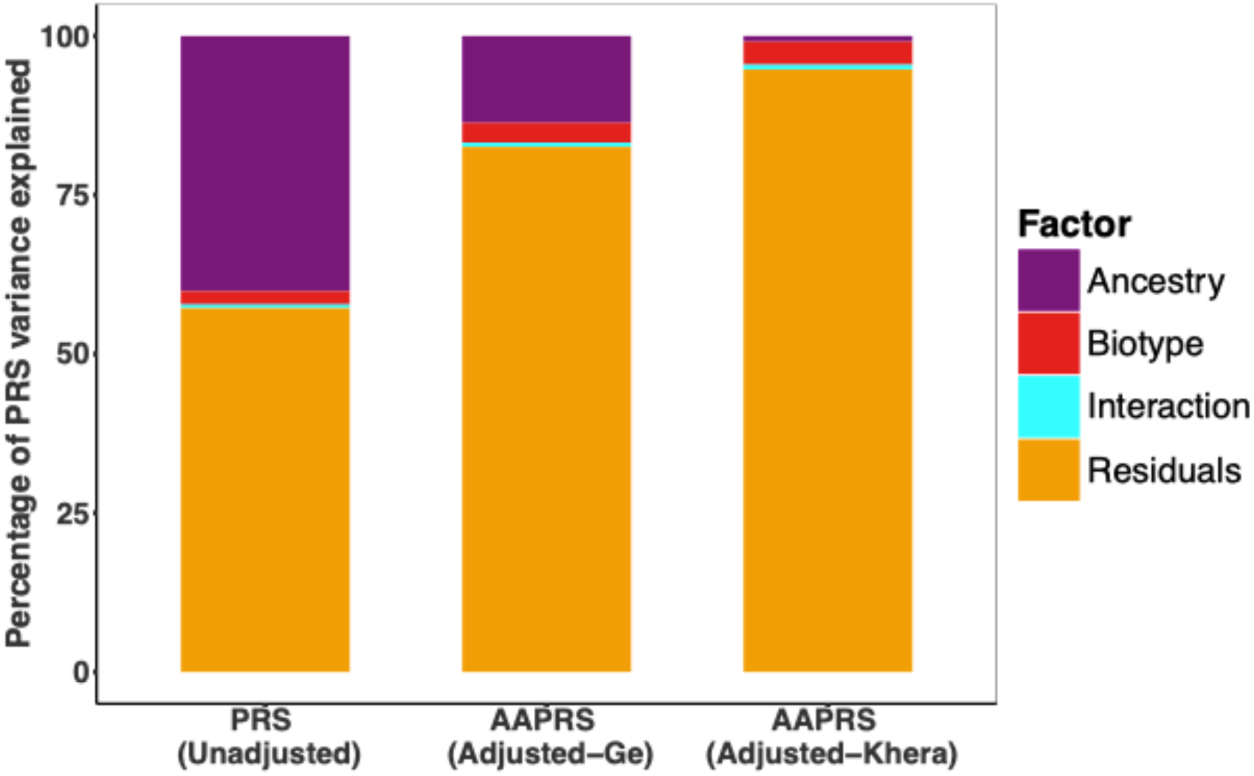
Percentage of SCZ AAPRS variance explained by each factor in two-way Analysis of Variance (ANOVA). “Unadjusted” is PRS-CSx *meta* PRS before *post hoc* ancestry adjustment. “Adjusted” refers to Ancestry-Adjusted Polygenic Risk Scores (AAPRSs) with *post hoc* ancestry adjustment of Khera or Ge. Minimal ancestry variance is desirable for AAPRS in a combined multi-ancestry sample. The residual variance would be the effect of SNPs on the PRS. Ideally, this would take up the largest share of the PRS variance, and the effects of the other variables would be minimized, as seen in the Khera AAPRS. With the Khera method, ancestry accounted for 1% of AAPRS variance (P = 4.37e-04) and Biotypes significantly accounted for 4% (P = 2.20e-17) vs. 14% (P = 3.03e-70) and 3% (P = 3.08e-17) for Ge. There were no significant interactions between Biotype and ancestry in either adjustment method (Table S6).

#### No overfitting observed with the Khera ancestry adjustment method

The question of overfitting in the Khera method is raised by its use of its own healthy controls for training its linear regression model. To address this question, we determined whether it matters if the training group is inside the total sample or not. For this question, we did two-fold cross-validation, where the data was randomly divided into two equal parts, called “folds”, and used for training and testing separately.

With two-fold cross-validation, the estimated overlap of the Khera AAPRS density kernels among the five ancestries was 0.969 (95% CI: 0.960, 0.978). No significant overfitting was observed compared to the initial overlap statistic of Khera AAPRS 0.974 (95% CI: 0.966, 0.982).

#### The three Biotypes show no significant differences in SCZ AAPRSs

Wilcoxon tests showed significant differences between each Biotype and healthy controls in SCZ AAPRSs in the combined multi-ancestry sample, but no significant differences among the three Biotypes (Figure 5, Table S7).

**Figure 5.**
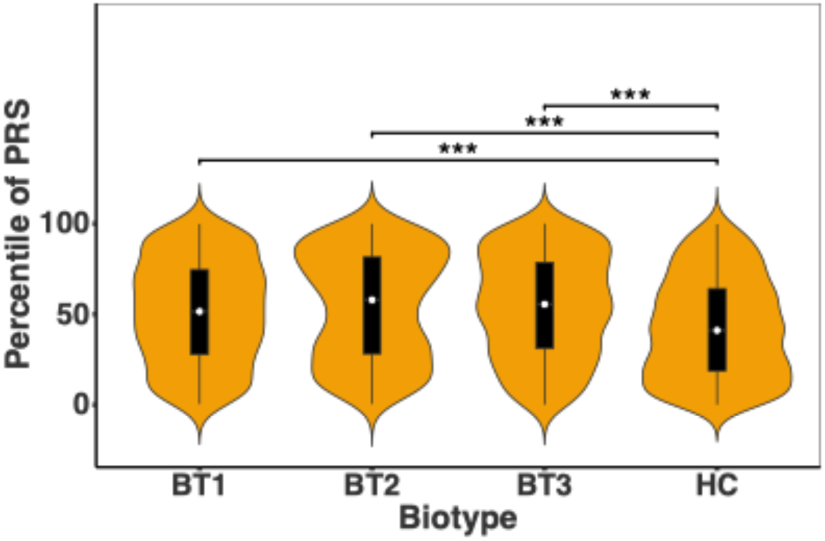
Biotype differences on SCZ AAPRS. AAPRS refers to Ancestry-Adjusted Polygenic Risk Score with *post hoc* ancestry adjustment of Khera. The differences of AAPRS among Biotypes in the combined multi-ancestry dataset are shown by violin plot. Wilcoxon tests were used for the comparison. Bonferroni-corrected significance threshold over 6 two-sample Wilcoxon tests is P-value < 8.33e-03. Only significant comparison results are labeled with asterisks. *** indicates P-value < 1.67e-04.

#### SCZ- and BD-GWAS summary statistics give similar AAPRSs across Biotypes within EUR ancestry

To see whether these three psychosis Biotypes share a polygenic risk for both SCZ and BD, we calculated PRS and AAPRS for EUR individuals in B-SNIP dataset using PGC EUR GWAS summary statistics from two different diagnoses, SCZ and BD. We studied only EUR ancestry B-SNIP participants because ancestry-specific BD GWAS summary statistics are currently available only for EUR ancestry.

Cross-diagnosis and within-diagnosis case-control prediction accuracy of unadjusted PRS and AAPRS was first evaluated by AUC and Nagelkerke’s pseudo-R^2^, as described in *Materials and methods*. The SCZ AAPRS performed slightly better than BD AAPRS on both Nagelkerke’s Pseudo-R^2^ and AUC in the combined diagnosis sample. SCZ AAPRS had good prediction accuracy for cross-diagnosis case-control status (that is, for BD persons). However, BD AAPRS had less cross-diagnosis prediction accuracy (that is, for SCZ persons). Integrating the two sets of GWAS summary statistics (EUR SCZ and EUR BD) into a combined psychosis AAPRS gave better prediction accuracy of case-control status for BD persons and the most consistently improved prediction accuracy for the combined diagnoses (BD and SCZ persons) (Figure 6).

**Figure 6.**
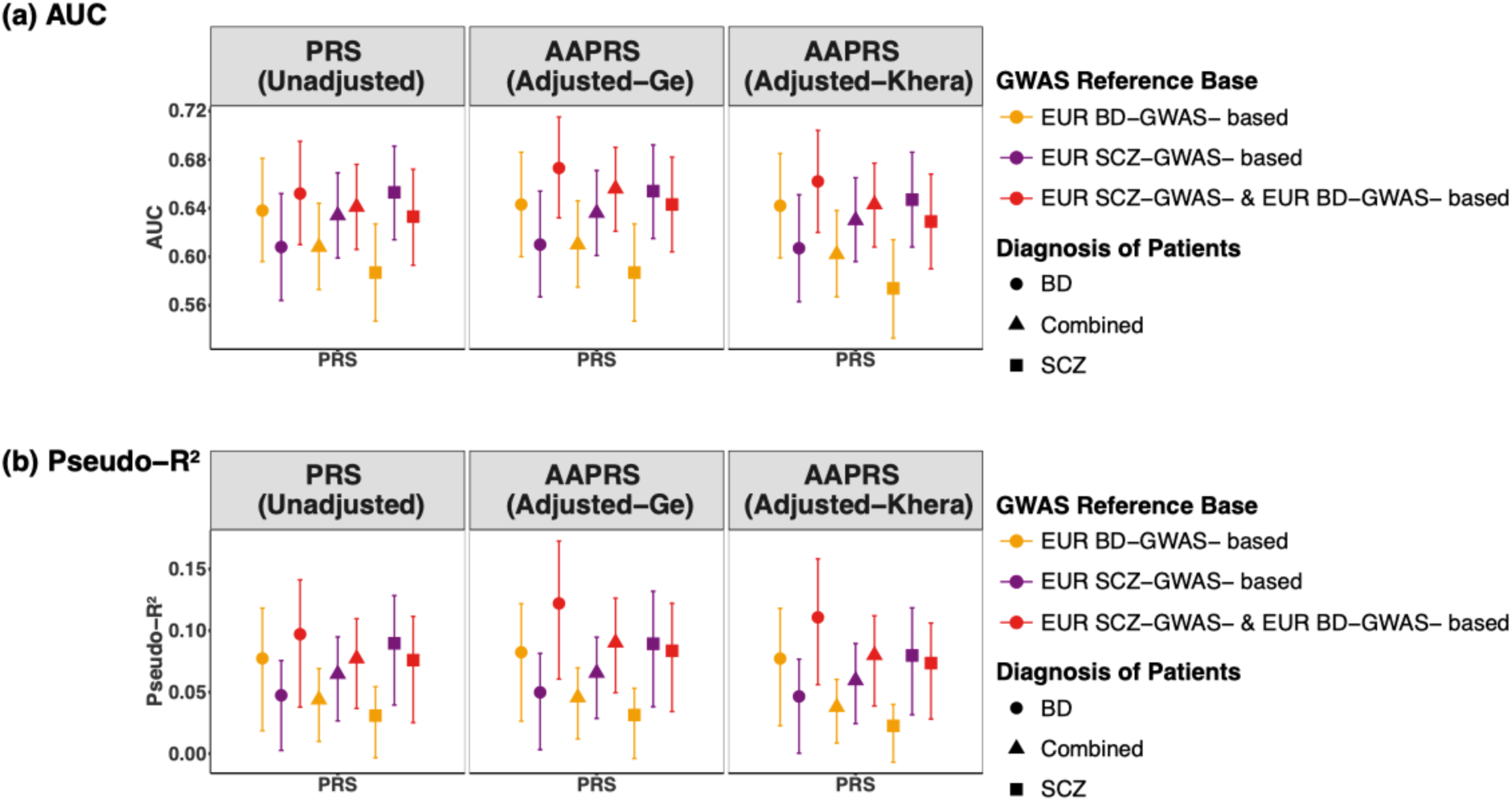
Prediction accuracy of EUR SCZ and BD GWAS-summary-statistics-based PRSs for case-control status for different diagnostic groups within EUR ancestry before and after ancestry adjustment. (a) The area under the receiver operating characteristic (ROC) curve (AUC) of PRSs. (b) The proportion of the case-control variance (Nagelkerke’s pseudo-R^2^) explained by PRSs. Lines for Nagelkerke’s pseudo-R^2^ in (b) correspond to 95% confidence intervals calculated via 1000 bootstrapping. PGC EUR SCZ and EUR BD GWAS summary statistics were used for PRS construction. “based” refers to the diagnosis of the GWAS summary statistics used to generate the PRS.

Two-sample Wilcoxon tests were used to determine whether there are differences among the three distinct Biotypes on both SCZ AAPRSs and BD AAPRSs within EUR ancestry. Results showed no significant differences among the three Biotypes, but each of the three Biotypes exhibited significant differences from healthy controls (Wilcoxon test, Bonferroni-corrected P < 8.33e-03) in the combined diagnostic persons in SCZ AAPRSs (Figure S3a), BD AAPRSs (Figure S3b), or psychosis AAPRSs (Figure S3c). In other words, the three psychosis Biotypes shared a polygenic risk based on both SCZ and BD GWAS summary statistics.

### 2. Molecular Associations

#### TWASs on imputed adult and fetal brain gene expression, mRNA isoforms, and RNA splicing identify 12 associations with Biotypes

We first evaluated overall case-control, SCZ-control, BD-control, SCZ-SAD, SCZ-BD, and SAD-BD associations in TWASs (gene-expression in both adult and fetal brain, isoforms in adult brain, and splicing events in adult brain), which would be broader than specific Biotype associations. However, no significant associations were detected.

We proceeded to perform separate TWAS analyses of each Biotype, comparing them with each other and with healthy controls. Twelve gene and isoform associations with Biotypes in adult or fetal brain were detected with multiple testing corrections (BH FDR < 0.05) applied (Table 1). No significant splicing event associations with Biotypes were detected.

**Table 1.** Imputed gene expression and mRNA isoforms associated with Biotypes.

| Gene Id | Gene Name | Comparison | Developmental Stage | Transcriptome | Ancestry | Effect Size | SE | Z-score | P-value | FDR |
| --- | --- | --- | --- | --- | --- | --- | --- | --- | --- | --- |
| ENSG00000165506 | DNAAF2 | HC-BT2 | adult | gene-level | SAS | 5.72 | 0.89 | 6.46 | 3.32E-07 | 0.005 |
| ENST00000608819 | ENSG00000272941, Novel Transcript, antisense to C7orf49 | HC-BT3 | adult | isoform-level | AFR | -0.95 | 0.18 | -5.28 | 2.40E-07 | 0.008 |
| ENST00000481410 | CYREN | HC-BT3 | adult | isoform-level | AFR | -0.89 | 0.18 | -4.95 | 1.23E-06 | 0.016 |
| ENST00000487774 | CYREN | HC-BT3 | adult | isoform-level | AFR | -0.88 | 0.18 | -4.83 | 2.12E-06 | 0.016 |
| ENST00000466307 | TMEM140 | HC-BT3 | adult | isoform-level | AFR | -1.08 | 0.22 | -4.84 | 2.05E-06 | 0.016 |
| ENST00000275767 | TMEM140 | HC-BT3 | adult | isoform-level | AFR | -0.59 | 0.12 | -4.82 | 2.28E-06 | 0.016 |
| ENST00000670978 | ENSG00000287733, Novel Transcript | HC-BT3 | adult | isoform-level | AFR | 0.29 | 0.06 | 4.71 | 3.64E-06 | 0.021 |
| ENSG00000257176 | Novel Transcript, antisense to FAR2 | HC-BT3 | fetal | gene-level | AFR | 0.74 | 0.16 | 4.75 | 3.08E-06 | 0.022 |
| ENSG00000162817 | C1orf115 | HC-BT1 | adult | gene-level | SAS | -4.04 | 0.72 | -5.63 | 2.38E-06 | 0.034 |
| ENST00000414809 | ARTN | HC-BT2 | adult | isoform-level | SAS | 1.87 | 0.31 | 6.01 | 1.18E-06 | 0.04 |
| ENSG00000287733 | Novel Transcript | HC-BT3 | fetal | gene-level | AFR | 1.03 | 0.23 | 4.45 | 1.19E-05 | 0.042 |
| ENSG00000275476 | Novel Transcript, antisense to FAR2 | HC-BT3 | fetal | gene-level | AFR | 2.13 | 0.49 | 4.33 | 1.97E-05 | 0.046 |

#### Mendelian Randomization (MR) detected seven putatively causal genes of Biotypes

Based on the twelve significantly Biotype-associated genes and transcripts (nine unique genes), we did a causal (mediation) exploration using MR-JTI. With Bonferroni correction, seven unique genes (four genes *TMEM140*, *ARTN*, *C1orf115*, *CYREN*, and three transcripts ENSG00000272941, ENSG00000257176, ENSG00000287733) showed significant potential causal relationships with psychosis Biotypes (Table 2).

**Table 2.**
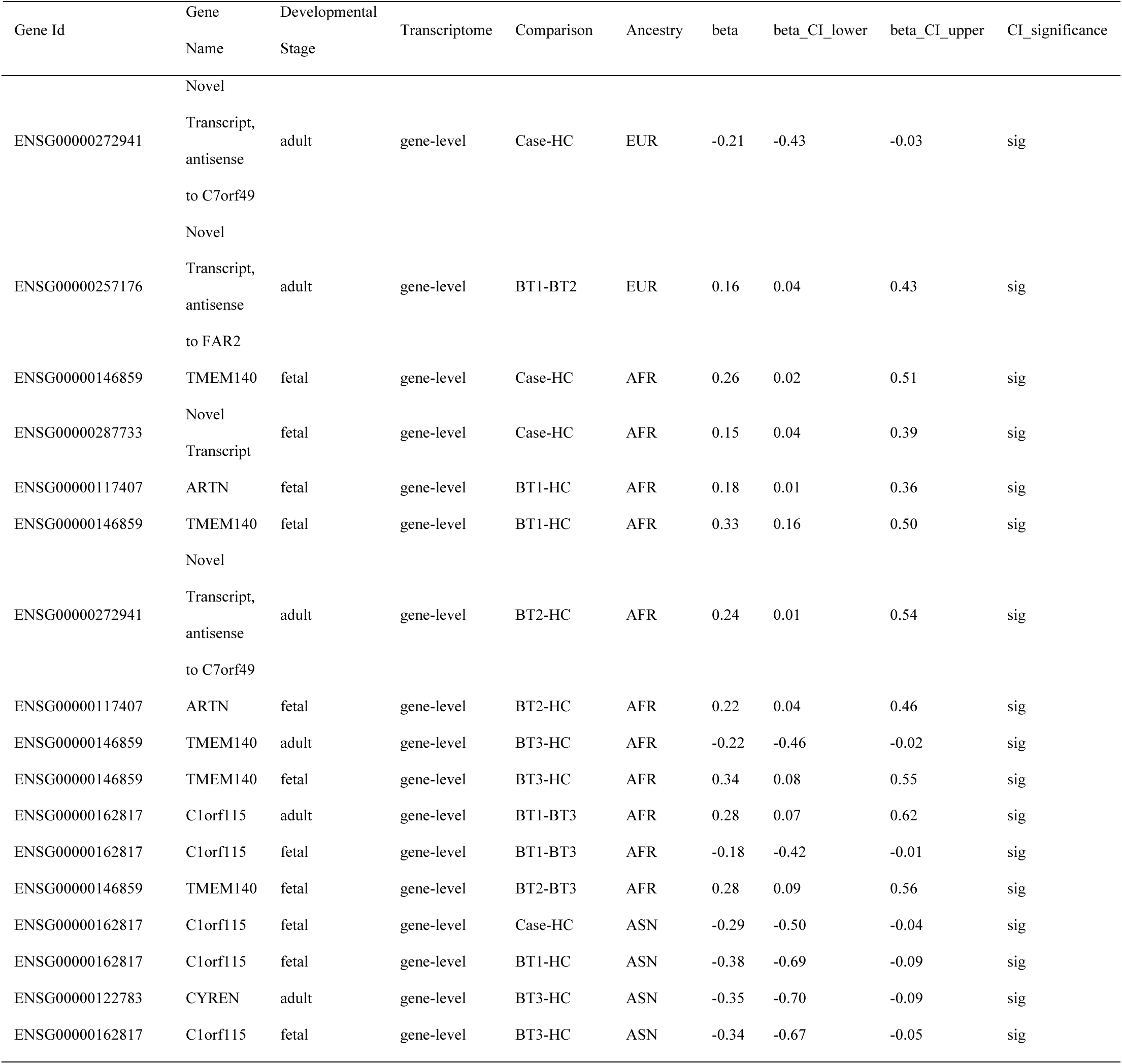
MR-JTI significant (Bonferroni corrected) results of the candidate genes/transcripts in Table 1.

The seven putatively causal genes of psychosis Biotypes are enriched (P < 0.05) in the biological pathways of Rearranged during Transfection (RET) signaling, Neural Cell Adhesion Molecule 1 (NCAM1) interactions, and NCAM signaling for neurite out-growth (Figure 7).

**Figure 7.**
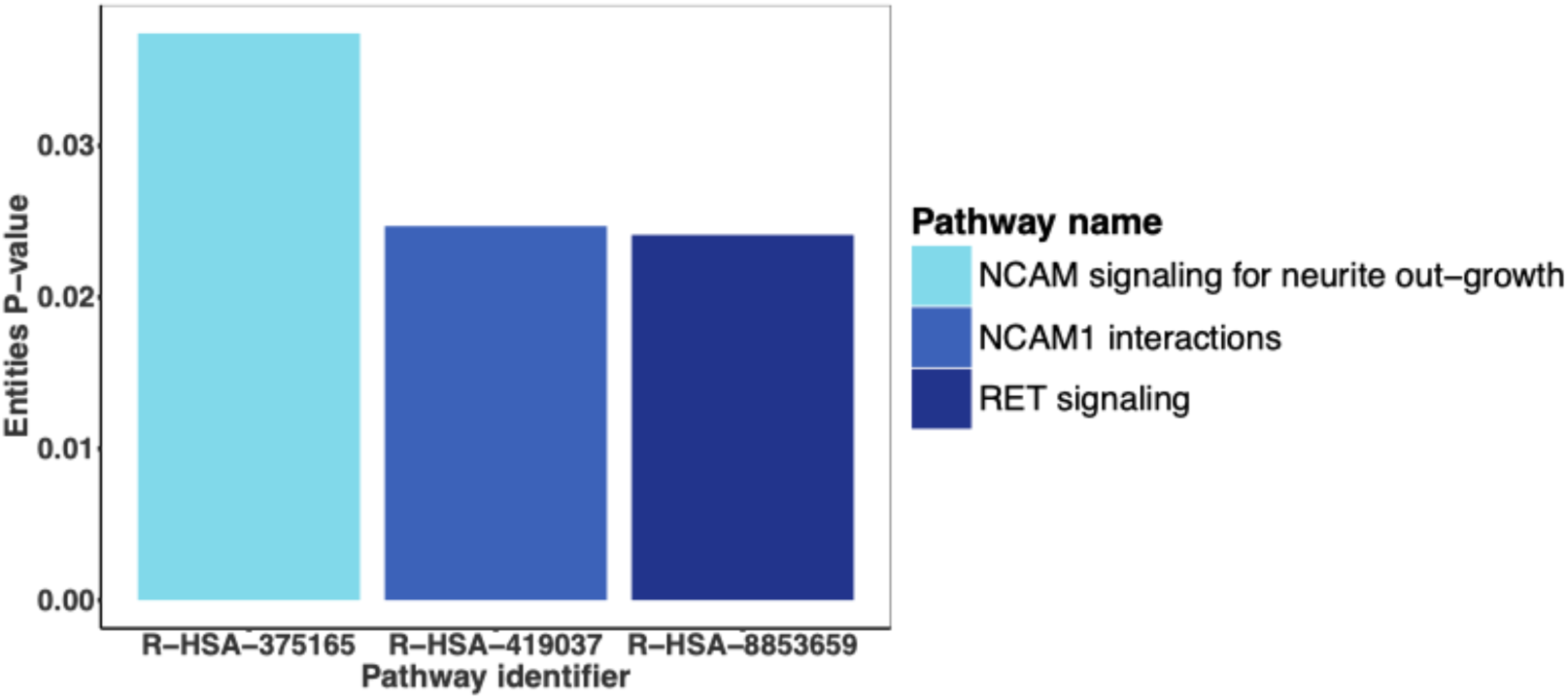
Histogram of significantly (P < 0.05) enriched biological pathways for the seven Biotype causal genes in Table 2. Legend: RET = Rearranged during Transfection, NCAM = Neural Cell Adhesion Molecule.

## Discussion

Non-portability of PRSs across different ancestries is well-known^44–50, 52–54, 88^. To be useful for multi-ancestry medical care, an AAPRS must satisfy requirements of accuracy and portability.

*Post-hoc* ancestry adjustments in multi-ancestry samples have recently been proposed to satisfy these requirements, as an alternative to meta-analysis by ancestry. We evaluated two methods of *post hoc* ancestry adjustment of PRS^52, 53^. In our comparative analysis in the B-SNIP dataset, the Khera method^52^ adjustment had better portability among ancestries, and the two methods had comparable accuracy. We also examined possible overfitting in the Khera method since part of the target data is used to fit the model to the entire target dataset and found no overfitting by cross-validation analysis. The Khera method was then chosen for analysis of the B-SNIP data, a multi-ancestry dataset which is the only existing dataset with psychosis Biotypes and genotypes.

The Biotypes are an innovation in diagnosis of psychosis disorders, as they are based on neurobiological measures as opposed to reported symptoms^35, 89^. In a previous study^35, 89^, Biotypes 1 and 2 had poorer scores on cognition and two other Biofactors (Intermediate Phenotypes) than Biotype 3 and healthy controls. On other Biofactors, Biotype 1 or Biotype 2 were more different from healthy controls than Biotype 3. We hypothesized that similar differences might exist in the overall polygenic risk of SCZ, with Biotype 3 closer to controls. However, we found no significant differences in AAPRSs among the three Biotypes, although each Biotype had higher AAPRS than healthy control, as expected. The PRS and AAPRS for the above analysis incorporated SCZ GWAS summary statistics from multiple ancestries using the PRS-CSx program. We also found that the Biotype AAPRS results were preserved in the polygenic risk of BD in EUR ancestry using EUR BD GWAS summary statistics. BD GWAS summary statistics from other ancestries were not available.

Interestingly, we found that in EUR ancestry, AAPRS based on SCZ GWAS summary statistics had comparable prediction accuracy for case-control status for BD persons, while AAPRS based on BD GWAS summary statistics had comparable (although smaller) prediction accuracy of case-control status for SCZ persons. Integrating the two sets of GWAS summary statistics (EUR SCZ and EUR BD) improved the prediction accuracy of case-control status for BD persons and for the combined diagnoses.

By projecting B-SNIP samples onto the 1KG PC space and using Random Forest (RF) to classify individuals into the five super populations, more genetically homogeneous groups can be identified for further analysis, while still retaining typically underrepresented groups. However, this approach is limited by the populations in the reference panel, which may not adequately represent certain samples or admixtures in the target data^90^. Specifically, the 1KG reference panel underrepresents admixed ancestry, as its AMR group predominantly reflects Latin American individuals, whose genetic profiles may not align with those of the admixed individuals in B-SNIP. As a result, RF may misclassify admixed individuals by favoring larger or more distinct ancestry groups.

In the data analyzed here, there is close correspondence between self-reported race and the three categories (EUR, AFR and ASN) derived by RF analysis, except for AMR ancestry, in which the concordance rate was only 16% (Table S1), due to the possible reasons mentioned above. The ancestry-specific TWAS results in this study are based on ancestry categories, where the AFR and EUR ancestry categories closely correspond to self-reported race. The number of EAS and SAS individuals is much smaller and gave us less confidence in those TWAS associations. No significant TWAS associations were found in AMR ancestry.

In the PRS analyses, since two alternative methods are available for PRS adjustment in multiple ancestries, we assessed the two methods. The Khera method is more successful than the Ge method in adjusting the PRS for effects of ancestry. Before adjustment, 40% of the PRS is attributable to ancestry by ANOVA analysis. After adjustment with the Ge method, there is still 14% variance attributable, but after adjustment with the Khera method there is 1% variance (Table S6). This difference may be due to the target sample projection onto the 1KG PC space in the Ge method, whereas the Khera method uses only the PCs of the actual sample. This illustrates another limitation of using the 1KG ancestries as a reference and supports the use of Khera method for PRS adjustment.

Additionally, PRS adjustment procedures did not completely erase the contributions of ancestry to AAPRS, although it was assumed that PRSs can be adjusted to have identical distributions across ancestries. Larger samples and optimized AAPRS methods to be developed may be needed to determine if ancestry effects can ever be eliminated. An alternative possibility exists, that ancestry-related differences exist in psychosis disorders that prevent identical PRS distributions across populations. For example, gene-environment interaction might limit PRS portability, which remains to be investigated.

Comprehensively delineating environment and gene-environment interactions in the estimation of psychiatric disease risk is a hefty task and is beyond the scope of the present work. However, it should be acknowledged that genetic ancestry differences are likely due to environmental influence. Environmental factors are numerous and themselves inter-related, and differentially so across ancestry groups. Ancestry groups overlap substantially with socially constructed racial group identities that tend to delineate disparities in how such factors are experienced.

Specifically, the experience is generally one of the disadvantages for those identifying/identified as Black or African American. The B-SNIP sample is comprised of mostly individuals self-identifying as White or Black/African American and living in the United States. It is likely these groups experienced differential healthcare system quality and access, stress associated with racism, intergenerational trauma, and even biases potentially contributing to diagnoses despite efforts at standardization, among many other interrelated factors. Socioeconomic status and other variables share substantial variance with racial group identity and delineate key impactful environmental experiences.

Genomic and genetic associations with Biotypes might be influenced by the socially environmental components, either as confounding variables or by gene-environment interactions. We cannot conclude that the observed PRS association with Biotypes is purely genetically determined. Social environmental components or their interactions with the genetic factors might contribute to the variation of Biotypes. These interactions regarding case-control differences could be investigated in future studies by adding socio-environmental variables to the linear models for analysis of genetic, genomic, and ancestry variation among Biotypes. This type of analysis was recently reported for MDD in Nepal, where demographic variables and environmental exposures explained a far greater proportion of variance in liability to lifetime MDD, while the depression PRS was not strongly associated with MDD^91^.

Inflation can exist in TWAS analysis mainly due to the polygenicity of the target trait, especially with a large sample size, as discussed recently^83–85^. We found some obvious inflation in TWAS results (lambda: 1.19 ∼ 3.6 in gene-level TWAS in adult brain, 1.08 ∼ 4.07 in gene-level TWAS in fetal brain, 1.16 ∼ 4.18 in isoform-level TWAS in adult brain, and 1.24 ∼ 5.16 in splicing-level TWAS in adult brain) in the combined multi-ancestry sample when we did not include genotype PCs in the association analysis. The inflation was successfully addressed (all lambdas close to 1) by including the first five genotype PCs as covariates in a logistic regression model. No inflation existed in the various ancestry-specific and combined multi-ancestry genotype PC-corrected TWAS results (Tables S8-S11). A representative Q-Q plot illustrated the inflation effect in the association analysis results with and without genotype PC correction (Figure 8). We applied genotype PC correction to all the TWAS results.

**Figure 8.**
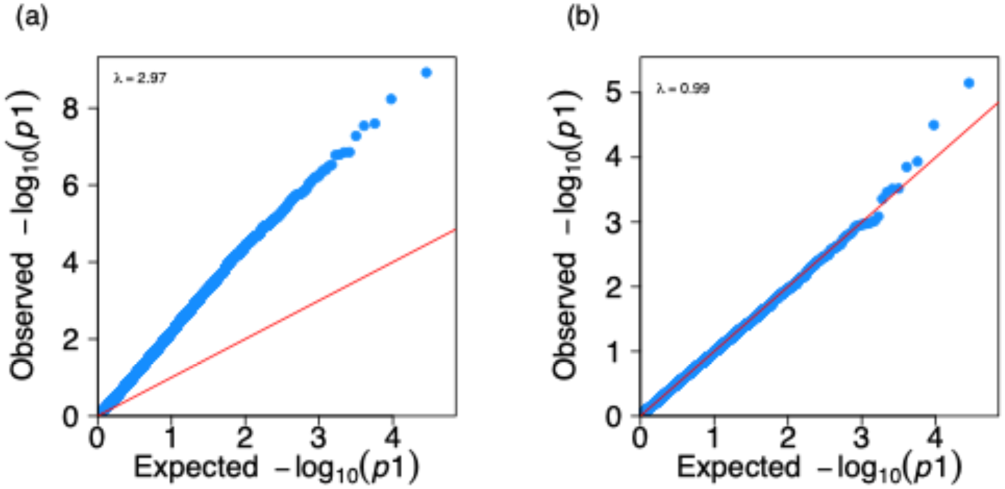
Representative Quantile-Quantile (Q-Q) plot of gene-level TWAS results in adult brain for Biotype 1 versus Healthy Control in the combined sample of the five ancestries. (a) QQ plot of TWAS results without covariates. (b) QQ plot of TWAS results with the first five genotype principal components (PCs) as covariates. Each blue dot represents for a gene. Results show that the inflation was successfully addressed by including the genotype PCs in the logistic regression model in the association test.

Twelve TWAS gene/isoform associations were found in Biotypes versus healthy control (Table 1). No significant associations were found in case-control, SCZ-control, BD-control, SCZ-SAD, SCZ-BD, or SAD-BD omics comparisons. This implies that association signals might be buried when using only traditional diagnosis, because there is genomic heterogeneity among Biotypes. Molecular, pharmacological, and genetic studies of Biotypes, as well as environmental factors, may be useful for disentangling at least part of the etiological and pathophysiological heterogeneity of psychosis.

Several Biotype-associated genes found in our study have been reported to be associated with psychosis disorders or neuropsychiatric disorders by previous genetic or genomic studies.

*C1orf115* was found to be differentially expressed when comparing SCZ, MDD, and ASD patients to healthy controls in multiple brain regions^92, 93^. *C1orf115* was also in GWAS associated (P = 3e-6) with response to antipsychotic therapy^94^. *DNAAF2* was found to be differentially expressed in BD and MDD frontal cortex^92^. *TMEM140* was reported as a down-regulated (logFC = -0.15, FDR q-value = 0.004) gene in first-episode psychosis patients in blood^95^. *TMEM140* is also associated with the prognosis of glioma by promoting cell viability and invasion^96^. *CYREN*, as known as *C7orf49*, was reported to be associated with Alzheimer’s disease (AD) and Parkinson’s disease (PD)^97^. *TMEM140* and *C7orf49* were reported to be associated with ADHD on the epigenomics^98^. *ARTN* was reported to be significantly associated (P = 1.6e-09) with SCZ and ADHD by meta-analyzing the common variant associations with SCZ and ADHD using Multi-marker Analysis of GenoMic Annotation (MAGMA)^99^. *ARTN* was reported to be down-regulated in patients with MDD in a current depressive state in blood^100^.

Seven of the twelve associated genes and isoforms are putative causal genes of Biotypes based on MR analysis, although causality here has the limitations associated with MR analyses, which do not account for complex interactions with other factors contributing to psychosis illnesses.

Nonetheless, these observations are relevant. These seven genes are enriched in psychosis and neuropsychiatric disorder-related biological pathways via RET signaling, NCAM1 interactions, and NCAM signaling for neurite out-growth (Figure 7). RET signaling is activated by glial cell line–derived neurotrophic factor (GDNF) ligands^101^. RET signaling was reported to function in motor neurons^102^, including dopaminergic (DA) neurons^103^, and associated with various diseases, such as PD^104^. It was also reported to be associated with vitamin D deficiency which increases the risk of SCZ^105^. The NCAM was reported to play important roles in neurite outgrowth, long-term potentiation in the hippocampus and synaptic plasticity^106^ in both the developing and adult brains^107^. As a synaptic plasticity marker, NCAM1 was reported to be differentially altered in SCZ^108, 109^ and BD^110^ patients, and the SNPs within NCAM1 contribute to the risk of both SCZ and BD possibly through alternative splicing of the gene^110^. The NCAM1 gene set was also reported to be linked to depressive symptoms^111^.

To validate the three biological pathways in Figure 7 and suggest additional pathways, we relaxed the TWAS significance threshold to Benjamini & Hochberg (BH) FDR < 0.1, thus including 11 additional suggestive gene/transcript/splicing associations with Biotypes (Table S12), besides the 12 significant associations in Table 1. We then did the causal analysis for all the 23 associations (19 unique genes) using MR-JTI program, and 14 unique genes are suggestively causal genes (because of the relaxed TWAS significance threshold) of Biotypes or psychosis disorders. These 14 suggestively causal genes are enriched in 31 additional biological pathways using a threshold BH FDR < 0.1 (Table S13), and in the three biological pathways shown in the Main text Figure 7. Some of these additional biological pathways, such as Nonsense Mediated Decay (NMD) related pathways^112, 113^, response of EIF2AK4 (GCN2) to amino acid deficiency^114^, signaling by NTRK3 (TRKC)^115–117^, axon guidance^118^, and nervous system development^119^, are reported to be therapeutic targets or play important roles in psychosis disorders.

To interrogate the robustness of PRS and TWAS findings, we performed analyses based on resampling ten times, since no other data of Biotypes can be used for replication. Resampling results are consistent showing that the Khera method provided better portability and comparative accuracy in SCZ AAPRS, and the three Biotypes share a polygenic risk of psychosis disorders. Resampling also showed the TWAS results of eleven genes and transcripts (except for ENSG00000287733) replicated 1 to 4 times (Table S14).

Despite the resampling results, we note that gene associations with psychosis Biotypes found in samples of Asian (EAS or SAS) ancestry have a much smaller sample size than the other ancestries. These results, although statistically significant, are offered as tentative conclusions. The three associated genes found in SAS ancestry would require further support. We did not find any genes that are significantly associated with EAS ancestry. In short, the genes associated with Biotypes in Asian ancestry should be viewed with caution.

In the PRS analyses, including or excluding Asian ancestry did not significantly affect SCZ prediction accuracy, ancestry adjustments, or Biotype comparisons. There was no significant change in prediction performance or ancestry adjustment results with or without Asian SCZ GWAS data or Asian samples from the target dataset (B-SNIP) (Figures 2, S4). Additionally, no significant differences in SCZ AAPRSs were found among the three Biotypes, regardless of whether including or excluding Asian SCZ GWAS data or Asian samples from the target dataset data (B-SNIP) (Table S7).

The eQTL, sQTL, and isoQTL models used for TWAS analyses in this study are mainly based on EUR samples. This might cause lowered prediction accuracy of GReX components for other ancestry individuals in the B-SNIP dataset, resulting in low power of detecting associations in other ancestries.

In conclusion, our results suggest that the three psychosis Biotypes share comparable polygenic risks based on PRS calculated from both SCZ and BD GWAS summary statistics. Furthermore, different Biotypes have specific psychosis risk genes, some of which are putatively causal genes with specific biological pathways. The molecular associations with Biotypes in this research suggest that pharmacological clinical trials and biological investigations might benefit from analyzing results separately by Biotypes as well as the usual analysis by diagnosis. That is, Biotypes may become a component of personalized diagnosis and treatment.

## Data availability

B-SNIP data could be obtained from the NIMH Data Archive (https://nda.nih.gov; NDAR ID: 2274, respectively). The 1000 Genomes phase 3 genotype data are available at https://www.cog-genomics.org/plink/2.0/resources#phase3_1kg. Other data sources used in this study are provided in the *Materials and methods* section and Tables S3-S4.

## Code availability

PRS-CSx implementation script is available at https://github.com/getian107/PRScsx. PRS-CS implementation script is available at https://github.com/getian107/PRScs. Ancestry assignment script is available at https://github.com/Annefeng/PBK-QC-pipeline. PLINK 2.0 is available at https://www.cog-genomics.org/plink/2.0/.

## Acknowledgments

This research was supported by several NIH grants: Tamminga: MH096913, MH077851 Pearlson: MH096957, MH077945 Keshavan: MH096942, MH078113 Keedy: MH103368, MH124804, MH127162 Clementz: MH124803, MH126398, MH096900, MH124806, MH103366, MH124802, MH127172 Parker: NIH/NIMH R01MH117315 and NIH/NCATS UL1TR002378, TL1TR002382 Liu: U01 MH122591, 1U01MH116489, 1R01MH110920, R01MH126459 Gershon: MH103368, MH124804, MH127162

This research was also supported by the National Natural Science Foundation of China (Grants Nos. 82022024), and the science and technology innovation Program of Hunan Province (2021RC4018, 2021RC5027). Innovation-driven Project of Central South University (Grant No. 2020CX003).

The authors gratefully acknowledge support from the Christopher Eklund family and the Geraldi Norton Foundation. We thank Drs Tian Ge and Hailiang Huang for their correspondence and support about PRS *post hoc* ancestry adjustment. This work was completed in part with resources provided by the University of Chicago’s Research Computing Center.

## Author contributions

ESG, CL and CX conceived and designed the study. ESG, CL and CC supervised the study and helped interpreting of data. CX did the formal analyses and drafted the manuscript. NAR did the B-SNIP genotype preprocessing (imputation and quality control) analysis. CW and RD built the PsychENCODE fetal brain predict.db. ESG, CL, CC, CX, NAR, CAT, MSK, GDP, SKK, BC, JEM, DP, RL, SKH, JRB, EII, CW, and RD reviewed, edited, and approved the final version of the manuscript.

## Competing interests

Cuihua Xia: None.

Ney Alliey-Rodriguez: None.

Carol A. Tamminga: B-SNIP Diagnostics, Board of Managers; Kynexis, Scientific Advisory Board and retainer; Merck DSMB; Neuventis, Board, own stock.

Matcheri S. Keshavan: B-SNIP Diagnostics, Board of Managers; Advisor to Alkermes. Godfrey D. Pearlson: B-SNIP Diagnostics, Board of Managers.

Sarah K. Keedy: B-SNIP Diagnostics, Board of Managers.

Brett A. Clementz: B-SNIP Diagnostics, Board of Managers; Kynexis Corporation, Scientific Advisory Board.

Jennifer E. McDowell: B-SNIP Diagnostics, Board of Managers. David A. Parker: None.

Rebekka Lencer: None.

S. Kristian Hill: None.

Jeffrey R. Bishop: None.

Elena I. Ivleva: B-SNIP Diagnostics, Board of Managers. Cindy Wen: None.

Rujia Dai: None. Chao Chen: None. Chunyu Liu: None.

Elliot S. Gershon: B-SNIP Diagnostics, Board of Managers; Consultant: Kynexis Corporation.

## Ethics approval and consent to participate

All methods were performed in accordance with the relevant guidelines and regulations. B-SNIP recruitment sites were in Athens GA (University of Georgia and Augusta University Medical College of Georgia), Baltimore MD (Maryland Psychiatric Research Center), Boston MA (Beth Israel Deaconess Medical Center), Chicago IL (University of Illinois-Chicago and University of Chicago), Dallas TX (UT Southwestern Medical Center), Detroit MI (Wayne State University), and Hartford CT (Institute of Living). All recruitments, interviews, and laboratory data collections were completed at those locations. The Institutional Review Board at participating institutions approved the projects; participants provided informed consent prior to involvement.

