## Supplementary Information for "Genetic Analysis of Psychosis Biotypes: Shared Ancestry-Adjusted Polygenic Risk and Unique Genomic Associations"

### Table of contents

**Summary of Supplementary Information**

**Supplementary Tables**

Table S1. Concordance between 1000 Genomes Random Forest (RF) inferred ancestry and Self-reported race questionnaire.

Table S2. Number of individuals in each Biotype by Ancestry.

Table S3. GWAS summary statistics used in this study.

Table S4. xQTL databases (PredictDB) used in this study.

Table S5. Prediction accuracy of SCZ PRS for case-control status under different ancestry adjustment methods.

Table S6. Two-way Analysis of Variance (ANOVA) of Ancestry and Biotype effects on Schizophrenia PRS under different ancestry adjustment methods.

Table S7. Wilcoxon test statistics of Biotype differences on percentile-transformed SCZ AAPRSs.

Table S8. TWAS of adult brain gene expression: Inflation of results with and without genotype principal components (PCs) as covariates.

Table S9. TWAS of fetal brain gene expression: Inflation of results with and without genotype principal components (PCs) as covariates.

Table S10. TWAS of adult brain isoform expression: Inflation of results with and without genotype principal components (PCs) as covariates.

Table S11. TWAS of adult brain splicing expression: Inflation of results with and without genotype principal components (PCs) as covariates.

Table S12. TWAS suggestive gene/transcript/splicing associations with the threshold FDR < 0.1.

Table S13. Enriched biological pathways of the 14 unique causal genes based on suggestive TWAS associations in Table S12.

Table S14. Resampling analysis for TWAS findings.

**Supplementary Figures**

Figure S1. B-SNIP individuals (N = 2,505) projected in 1KG Principal Components (PCs) 1 and 2 graphic space.

Figure S2. Principle of Mendelian Randomization (from Thomas and Conti).

Figure S3. Biotype vs. healthy control and Biotype vs. Biotype differences in AAPRSs based on SCZ-, BD- or both SCZ- and BD- GWAS summary statistics within EUR ancestry B-SNIP participants.

Figure S4. Prediction accuracy of PRSs for case-control status before and after ancestry adjustment within and across 5 ancestries based on different GWAS summary statistics.

**References**

#### Summary of Supplementary Information

This document summarizes the ancestry information (Table S1, Figure S1), sample size in each Biotype by ancestry (Table S2), data sources (Tables S3-S4), prediction accuracy of SCZ PRS for case-control status (Table S5), PRS variance explained by ancestry (Table S6), test statistics of Biotype differences on percentile-transformed SCZ AAPRSs (Table S7), inflation of TWAS results (Tables S8-S11), suggestive TWAS associations (Table S12), enriched biological pathways of the 14 unique causal genes based on suggestive TWAS associations (Table S13), resampling analysis for TWAS findings (Table S14), principle of Mendelian Randomization (Figure S2), Biotype comparison results on PRS within EUR individuals in B-SNIP dataset (Figure S3), and prediction accuracy of PRSs for case-control status before and after ancestry adjustment within and across 5 ancestries based on different GWAS summary statistics (Figure S4).

Note on abbreviations in Supplementary Figures and Tables: EUR = European, AFR = African, AMR = Admixed American, EAS = East Asian, SAS = South Asian. Diagnoses (as defined in article text): SCZ = Schizophrenia, BD = Bipolar Disorder.

### Supplementary Tables

**Table S1. Concordance between 1000 Genomes Random Forest (RF) inferred ancestry and Self-reported race questionnaire.**

| **RF-inferred ancestry** | **Self-reported race** | | | | | | | |
|---|---|---|---|---|---|---|---|---|
| | **AA** | **AE** | **AS** | **CA** | **MR** | **NH** | **OT** | **UNK** |
| **AFR** | 677 | 3 | 1 | 4 | 34 | 0 | 8 | 1 |
| **AMR** | 3 | 7 | 1 | 123 | 14 | 0 | 34 | 3 |
| **EAS** | 1 | 0 | 40 | 6 | 11 | 2 | 1 | 1 |
| **EUR** | 13 | 1 | 3 | 859 | 29 | 0 | 12 | 0 |
| **SAS** | 2 | 0 | 41 | 0 | 0 | 0 | 1 | 0 |

Legend: Self-reported race: response of each person based on current cultural categories. AA = African American, AE = American Indian, AS = Asian, CA = Caucasian, MR = Multiracial/Mixed Race, NH = Native Hawaiian, OT = Other Race, UNK = Unknown/Missing. A Random-Forest (RF) method was used to assign each person in the B-SNIP dataset to one of the five 1000 Genomes (1KG) super populations – European (EUR), African (AFR), Admixed American (AMR), East Asian (EAS) and South Asian (SAS). Within ancestry, the concordance rates of RF-inferred ancestry with self-reported race in EUR, AFR, ASN (Asian: EAS + SAS) and AMR are 87%, 97%, 94% and 16%, respectively.

**Table S2. Number of individuals in each Biotype by Ancestry.**

| Biotype | AFR | AMR | EAS | EUR | SAS |
|---|---|---|---|---|---|
| BT1 | 256 (44%) | 47 (34%) | 13 (38%) | 172 (24%) | 7 (37%) |
| BT2 | 213 (36%) | 41 (30%) | 7 (21%) | 219 (31%) | 3 (16%) |
| BT3 | 119 (20%) | 49 (36%) | 14 (41%) | 324 (45%) | 9 (47%) |
| HC | 200 | 75 | 35 | 345 | 30 |

Legend: EUR = European, AFR = African, AMR = Admixed American, EAS = East Asian, SAS = South Asian. BT1 = Biotype 1, BT2 = Biotype 2, BT3 = Biotype 3. The percentages displayed in each column represent the proportion of a specific Biotype relative to the total count of Biotypes within the corresponding ancestry.

#### Table S3. GWAS summary statistics used in this study.

| Disorder | Ancestry | Data Source | N_total | N_cases | N_controls | Filename | Source Link |
|---|---|---|---|---|---|---|---|
| SCZ | EUR | PGC 3 | 127,906 | 52,017 | 75,889 | PGC3_SCZ_wave3.european.autosome.public.v3.vcf.tsv.gz | https://figshare.com/articles/dataset/scz2022/19426775 |
| SCZ | AFR | PGC 3 | 9,824 | 5,998 | 3,826 | PGC3_SCZ_wave3.afram.autosome.public.v3.vcf.tsv.gz | https://figshare.com/articles/dataset/scz2022/19426775 |
| SCZ | ASN | PGC 3 | 27,363 | 12,305 | 15,058 | PGC3_SCZ_wave3.asian.autosome.public.v3.vcf.tsv.gz | https://figshare.com/articles/dataset/scz2022/19426775 |
| BD | EUR | PGC | 413,466 | 41,917 | 371,549 | pgc-bip2021-all.vcf.tsv.gz | https://figshare.com/ndownloader/articles/14102594/versions/2 |

#### Table S4. xQTL databases (PredictDB) used in this study.

| Developmental Stage | Brain Region | xQTL Type | N_total | Data Source | Source Link |
|---|---|---|---|---|---|
| Adult | Cortex | eQTL | 1,866 | PsychENCODE1 | https://predictdb.org/post/2020/07/19/psychencode-brain-expression-models |

| | | | | | |
|---|---|---|---|---|---|
| Adult | Frontal Cortex (BA9) | sQTL | 156 | GTEx v8 (elastic net-based) | https://predictdb.org/post/2021/07/21/gtex-v8-models-on-eqtl-and-sqtl/ |
| Adult | Cortex | isoQTL | 2,365 | PsychENCODE1 & AMP-AD | https://zenodo.org/record/8048198 |
| Fetal | Cortex | eQTL | 672 | PsychENCODE2 | https://www.synapse.org/#!Synapse:syn51184101.1 |

**Table S5. Prediction accuracy of SCZ PRS for case-control status under different ancestry adjustment methods.**

| PRS | Ancestry | AUC (95% CI) | Pseudo-$R^2$ (95% CI) |
|---|---|---|---|
| PRS (Unadjusted) | ALL | 0.606 (0.580, 0.631) | 0.043 (0.021, 0.062) |
| AAPRS (Adjusted-Khera) | ALL | 0.607 (0.582, 0.632) | 0.044 (0.021, 0.063) |
| AAPRS (Adjusted-Ge) | ALL | 0.619 (0.594, 0.644) | 0.053 (0.028, 0.075) |
| PRS (Unadjusted) | AFR | 0.546 (0.501, 0.591) | 0.010 (-0.013, 0.020) |
| AAPRS (Adjusted-Khera) | AFR | 0.553 (0.508, 0.599) | 0.013 (-0.011, 0.025) |
| AAPRS (Adjusted-Ge) | AFR | 0.551 (0.506, 0.596) | 0.012 (-0.011, 0.023) |
| PRS (Unadjusted) | AMR | 0.601 (0.520, 0.682) | 0.040 (-0.045, 0.078) |
| AAPRS (Adjusted-Khera) | AMR | 0.640 (0.560, 0.719) | 0.064 (-0.041, 0.118) |
| AAPRS (Adjusted-Ge) | AMR | 0.611 (0.531, 0.692) | 0.046 (-0.034, 0.088) |
| PRS (Unadjusted) | EAS | 0.623 (0.488, 0.757) | 0.051 (-0.142, 0.101) |
| AAPRS (Adjusted-Khera) | EAS | 0.611 (0.476, 0.746) | 0.053 (-0.117, 0.105) |
| AAPRS (Adjusted-Ge) | EAS | 0.617 (0.482, 0.752) | 0.052 (-0.131, 0.103) |
| PRS (Unadjusted) | EUR | 0.647 (0.612, 0.681) | 0.080 (0.041, 0.112) |
| AAPRS (Adjusted-Khera) | EUR | 0.644 (0.609, 0.678) | 0.076 (0.034, 0.107) |

| | | | |
|---|---|---|---|
| AAPRS (Adjusted-Ge) | EUR | 0.650 (0.615, 0.684) | 0.083 (0.041, 0.115) |
| PRS (Unadjusted) | SAS | 0.567 (0.394, 0.740) | 0.014 (-0.176, 0.028) |
| AAPRS (Adjusted-Khera) | SAS | 0.596 (0.427, 0.766) | 0.042 (-0.183, 0.084) |
| AAPRS (Adjusted-Ge) | SAS | 0.612 (0.445, 0.780) | 0.045 (-0.175, 0.090) |

**Table S6. Two-way Analysis of Variance (ANOVA) of Ancestry and Biotype effects on Schizophrenia PRS under different ancestry adjustment methods.**

| PRS | Factor | Percentage of Variance Explained | F_value | Pr_F | Df | Sum_of_Squares | Mean_Square |
|---|---|---|---|---|---|---|---|
| PRS (unadjusted) | Ancestry | 40.14 | 378.59 | 3.28e-247 | 4 | 314.98 | 78.75 |
| | Biotype | 2.07 | 26.05 | 1.51e-16 | 3 | 16.25 | 5.42 |
| | Interaction | 0.59 | 1.86 | 0.03 | 12 | 4.64 | 0.39 |
| | Residuals | 57.20 | | | 2158 | 448.86 | 0.21 |
| AAPRS (adjusted-Khera) | Ancestry | 0.89 | 5.10 | 4.37e-04 | 4 | 3.99 | 1.00 |
| | Biotype | 3.61 | 27.39 | 2.20e-17 | 3 | 16.08 | 5.36 |
| | Interaction | 0.76 | 1.44 | 0.14 | 12 | 3.38 | 0.28 |
| | Residuals | 94.74 | | | 2158 | 422.26 | 0.20 |
| AAPRS (adjusted-Ge) | Ancestry | 13.65 | 89.21 | 3.03e-70 | 4 | 297.17 | 74.29 |
| | Biotype | 3.12 | 27.16 | 3.08e-17 | 3 | 67.85 | 22.62 |
| | Interaction | 0.68 | 1.48 | 0.13 | 12 | 14.76 | 1.23 |
| | Residuals | 82.55 | | | 2158 | 1797.22 | 0.83 |

Legend: Contribution of ancestry to PRS variance is smallest in the Khera method. Residuals include cumulative effects of individual SNPs.

**Table S7. Wilcoxon test statistics of Biotype differences on percentile-transformed SCZ AAPRSs.**

| GWAS Reference Base | Group1 | Group2 | N1 | N2 | Statistic | P-value |
|---|---|---|---|---|---|---|
| EUR/AFR/ASN SCZ-GWAS- based | BT1 | BT2 | 495 | 483 | 111376 | 0.064 |
| | BT1 | BT3 | 495 | 515 | 120689 | 0.144 |
| | BT2 | BT3 | 483 | 515 | 126417 | 0.653 |
| | BT1 | HC | 495 | 685 | 199549 | **2.05E-07** |
| | BT2 | HC | 483 | 685 | 204640 | **4.95E-12** |
| | BT3 | HC | 515 | 685 | 216619 | **1.28E-11** |
| EUR/AFR SCZ-GWAS- based | BT1 | BT2 | 495 | 483 | 113114 | 0.146 |
| | BT1 | BT3 | 495 | 515 | 122366 | 0.272 |
| | BT2 | BT3 | 483 | 515 | 126070 | 0.709 |
| | BT1 | HC | 495 | 685 | 205639 | **4.12E-10** |
| | BT2 | HC | 483 | 685 | 207840 | **7.99E-14** |
| | BT3 | HC | 515 | 685 | 219822 | **2.68E-13** |

Legend: AAPRSs used in the Wilcoxon tests refer to Ancestry-Adjusted Polygenic Risk Score with post hoc ancestry adjustment of Khera in the combined multi-ancestry dataset. Wilcoxon tests were used for the percentile-transformed SCZ AAPRS comparisons. Bonferroni-corrected significance threshold over 6 two-sample Wilcoxon tests is P-value < 8.33e-03. Only significant comparison results are highlighted with bold font.

**Table S8. TWAS of adult brain gene expression: Inflation of results with and without genotype principal components (PCs) as covariates.**

| Ancestry | Group | Brain Developmental Stage | Transcriptome | Lambda (noPCs) | Lambda (withPCs) | Inflation (noPCs) | Inflation (withPCs) |
|---|---|---|---|---|---|---|---|
| ALL | case-ctrl | adult | gene | 1.87 | 0.97 | 1.3 | 0.98 |
| ALL | HC-BT1 | adult | gene | 2.97 | 0.99 | 1.53 | 0.98 |
| ALL | HC-BT2 | adult | gene | 2.09 | 0.98 | 1.35 | 0.97 |
| ALL | HC-BT3 | adult | gene | 1.32 | 0.97 | 1.14 | 0.97 |
| ALL | BT1-BT2 | adult | gene | 1.18 | 0.97 | 1.06 | 0.97 |
| ALL | BT1-BT3 | adult | gene | 3.95 | 0.95 | 1.68 | 0.97 |
| ALL | BT2-BT3 | adult | gene | 2.64 | 0.96 | 1.49 | 0.97 |
| EUR | case-ctrl | adult | gene | 1.05 | 1.03 | 1 | 1 |
| EUR | HC-BT1 | adult | gene | 1.04 | 1.04 | 1 | 1 |
| EUR | HC-BT2 | adult | gene | 1 | 1 | 0.99 | 0.99 |
| EUR | HC-BT3 | adult | gene | 1.04 | 1.03 | 1 | 1 |

| | | | | | | | |
|---|---|---|---|---|---|---|---|
| EUR | BT1-BT2 | adult | gene | 1.02 | 1.02 | 0.99 | 0.99 |
| EUR | BT1-BT3 | adult | gene | 1.03 | 1.01 | 0.99 | 0.99 |
| EUR | BT2-BT3 | adult | gene | 1 | 1 | 0.97 | 0.98 |
| AFR | case-ctrl | adult | gene | 0.99 | 0.98 | 0.99 | 0.99 |
| AFR | HC-BT1 | adult | gene | 0.99 | 0.98 | 0.98 | 0.98 |
| AFR | HC-BT2 | adult | gene | 1.01 | 1.01 | 0.99 | 0.99 |
| AFR | HC-BT3 | adult | gene | 0.98 | 0.99 | 0.98 | 0.98 |
| AFR | BT1-BT2 | adult | gene | 0.98 | 0.96 | 0.98 | 0.98 |
| AFR | BT1-BT3 | adult | gene | 0.99 | 0.98 | 0.98 | 0.98 |
| AFR | BT2-BT3 | adult | gene | 0.97 | 0.97 | 0.98 | 0.98 |
| AMR | case-ctrl | adult | gene | 1.29 | 0.97 | 1.1 | 0.98 |
| AMR | HC-BT1 | adult | gene | 1.4 | 1 | 1.13 | 0.99 |
| AMR | HC-BT2 | adult | gene | 1.21 | 1.01 | 1.05 | 1.01 |
| AMR | HC-BT3 | adult | gene | 1.03 | 0.99 | 0.99 | 0.98 |
| AMR | BT1-BT2 | adult | gene | 1.07 | 1.03 | 0.99 | 1 |
| AMR | BT1-BT3 | adult | gene | 1.1 | 0.97 | 1.02 | 0.98 |
| AMR | BT2-BT3 | adult | gene | 1.04 | 0.97 | 0.98 | 0.99 |
| EAS | case-ctrl | adult | gene | 1 | 0.93 | 0.96 | 0.97 |
| EAS | HC-BT1 | adult | gene | 0.97 | 0.93 | 0.94 | 0.95 |
| EAS | HC-BT2 | adult | gene | 1.07 | 0.99 | 0.97 | 0.99 |
| EAS | HC-BT3 | adult | gene | 1.01 | 1.01 | 0.95 | 0.99 |
| EAS | BT1-BT2 | adult | gene | 1.04 | 0.91 | 0.92 | 0.97 |
| EAS | BT1-BT3 | adult | gene | 0.98 | 0.89 | 0.92 | 0.96 |
| EAS | BT2-BT3 | adult | gene | 1.05 | 0.93 | 0.92 | 0.97 |
| SAS | case-ctrl | adult | gene | 1.02 | 1 | 0.96 | 0.99 |
| SAS | HC-BT1 | adult | gene | 1.04 | 0.98 | 0.95 | 1 |
| SAS | HC-BT2 | adult | gene | 0.96 | 1 | 0.9 | 1.01 |
| SAS | HC-BT3 | adult | gene | 1.02 | 1.02 | 0.94 | 0.99 |
| SAS | BT1-BT2 | adult | gene | 0.96 | 1.04 | 0.8 | 1.01 |
| SAS | BT1-BT3 | adult | gene | 1.07 | 0.98 | 0.9 | 1.01 |
| SAS | BT2-BT3 | adult | gene | 0.95 | 1.03 | 0.83 | 1.04 |

Legend: PCs = Principal Components. “noPCs” means association results without any PCs included in the logistic regression model. “withPCs” means association results with the first five genotype PCs included in the logistic regression model. The metric *Inflation* is calculated using a Bayesian method based on the empirical null distribution. The metric *Lambda* is the genomic inflation factor, which is calculated by comparing the observed distribution of p-values to the expected uniform distribution under the null hypothesis of no association. A value close to 1 for both metrics indicates that the distribution of p-values closely follows the expected uniform distribution, suggesting minimal inflation. This result shows there is an inflation when genotype PCs are not adjusted for gene-level TWAS in adult brain in the combined multi-ancestry sample.

**Table S9. TWAS of fetal brain gene expression: Inflation of results with and without genotype principal components (PCs) as covariates.**

| Ancestry | Group | Brain Developmental Stage | Transcriptome | Lambda (noPCs) | Lambda (withPCs) | Inflation (noPCs) | Inflation (withPCs) |
|---|---|---|---|---|---|---|---|
| ALL | case-ctrl | fetal | gene | 1.74 | 1 | 1.27 | 0.98 |
| ALL | HC-BT1 | fetal | gene | 2.89 | 0.96 | 1.53 | 0.97 |
| ALL | HC-BT2 | fetal | gene | 1.91 | 1.03 | 1.32 | 0.97 |
| ALL | HC-BT3 | fetal | gene | 1.34 | 0.93 | 1.14 | 0.96 |
| ALL | BT1-BT2 | fetal | gene | 1.08 | 0.9 | 1.03 | 0.95 |
| ALL | BT1-BT3 | fetal | gene | 4.07 | 0.93 | 1.68 | 0.96 |
| ALL | BT2-BT3 | fetal | gene | 2.59 | 0.94 | 1.47 | 0.95 |
| EUR | case-ctrl | fetal | gene | 1.03 | 1 | 0.99 | 0.98 |
| EUR | HC-BT1 | fetal | gene | 1.02 | 1.03 | 0.98 | 0.98 |
| EUR | HC-BT2 | fetal | gene | 1.03 | 1.02 | 0.99 | 0.99 |
| EUR | HC-BT3 | fetal | gene | 0.95 | 0.95 | 0.98 | 0.98 |
| EUR | BT1-BT2 | fetal | gene | 1.03 | 1.05 | 0.98 | 0.99 |
| EUR | BT1-BT3 | fetal | gene | 1.04 | 1 | 0.99 | 0.99 |
| EUR | BT2-BT3 | fetal | gene | 0.98 | 0.96 | 0.97 | 0.97 |
| AFR | case-ctrl | fetal | gene | 1.06 | 1.06 | 1 | 1 |
| AFR | HC-BT1 | fetal | gene | 1.03 | 1.04 | 1 | 1 |
| AFR | HC-BT2 | fetal | gene | 1.02 | 1.01 | 0.99 | 0.99 |
| AFR | HC-BT3 | fetal | gene | 1.01 | 0.97 | 0.99 | 0.98 |
| AFR | BT1-BT2 | fetal | gene | 0.98 | 0.97 | 0.97 | 0.96 |
| AFR | BT1-BT3 | fetal | gene | 0.97 | 0.95 | 0.98 | 0.98 |
| AFR | BT2-BT3 | fetal | gene | 0.97 | 0.97 | 0.98 | 0.98 |

| | | | | | | | |
|---|---|---|---|---|---|---|---|
| AMR | case-ctrl | fetal | gene | 1.3 | 1 | 1.1 | 0.98 |
| AMR | HC-BT1 | fetal | gene | 1.45 | 1.02 | 1.14 | 1 |
| AMR | HC-BT2 | fetal | gene | 1.19 | 1 | 1.04 | 0.99 |
| AMR | HC-BT3 | fetal | gene | 1.09 | 0.99 | 0.99 | 0.98 |
| AMR | BT1-BT2 | fetal | gene | 1.08 | 1.03 | 1 | 1.01 |
| AMR | BT1-BT3 | fetal | gene | 1.13 | 0.97 | 1.03 | 0.98 |
| AMR | BT2-BT3 | fetal | gene | 1.1 | 1 | 0.98 | 0.98 |
| EAS | case-ctrl | fetal | gene | 1.01 | 0.94 | 0.97 | 0.97 |
| EAS | HC-BT1 | fetal | gene | 1.02 | 0.97 | 0.95 | 0.97 |
| EAS | HC-BT2 | fetal | gene | 1.11 | 1.02 | 0.97 | 0.99 |
| EAS | HC-BT3 | fetal | gene | 1.02 | 1 | 0.96 | 0.98 |
| EAS | BT1-BT2 | fetal | gene | 1.11 | 0.94 | 0.93 | 0.99 |
| EAS | BT1-BT3 | fetal | gene | 1.03 | 0.94 | 0.95 | 0.98 |
| EAS | BT2-BT3 | fetal | gene | 1.1 | 0.91 | 0.94 | 0.96 |
| SAS | case-ctrl | fetal | gene | 1.04 | 1.01 | 0.97 | 1 |
| SAS | HC-BT1 | fetal | gene | 1.06 | 0.99 | 0.95 | 1 |
| SAS | HC-BT2 | fetal | gene | 1.06 | 1.03 | 0.9 | 1.01 |
| SAS | HC-BT3 | fetal | gene | 1.06 | 1.02 | 0.95 | 1 |
| SAS | BT1-BT2 | fetal | gene | 1.05 | 1 | 0.83 | 1.01 |
| SAS | BT1-BT3 | fetal | gene | 1.06 | 0.98 | 0.92 | 1.02 |
| SAS | BT2-BT3 | fetal | gene | 1.12 | 1.02 | 0.86 | 1.04 |

Legend: PCs = Principal Components. “noPCs” means association results without any PCs included in the logistic regression model. “withPCs” means association results with the first five genotype PCs included in the logistic regression model. The metric *Inflation* is calculated using a Bayesian method based on the empirical null distribution. The metric *Lambda* is the genomic inflation factor, which is calculated by comparing the observed distribution of p-values to the expected uniform distribution under the null hypothesis of no association. A value close to 1 for both metrics indicates that the distribution of p-values closely follows the expected uniform distribution, suggesting minimal inflation. This result shows there is an inflation when genotype PCs are not adjusted for gene-level TWAS in fetal brain in the combined multi-ancestry sample.

**Table S10. TWAS of adult brain isoform expression: Inflation of results with and without genotype principal components (PCs) as covariates.**

| Ancestry | Group | Brain Developmental Stage | Transcriptome | Lambda (noPCs) | Lambda (withPCs) | Inflation (noPCs) | Inflation (withPCs) |
|---|---|---|---|---|---|---|---|
| ALL | case-ctrl | adult | isoform | 1.94 | 1 | 1.33 | 0.98 |
| ALL | HC-BT1 | adult | isoform | 3.13 | 0.99 | 1.61 | 0.98 |
| ALL | HC-BT2 | adult | isoform | 2.2 | 0.98 | 1.39 | 0.97 |
| ALL | HC-BT3 | adult | isoform | 1.4 | 0.94 | 1.15 | 0.97 |
| ALL | BT1-BT2 | adult | isoform | 1.16 | 0.94 | 1.06 | 0.96 |
| ALL | BT1-BT3 | adult | isoform | 4.18 | 0.99 | 1.8 | 0.98 |
| ALL | BT2-BT3 | adult | isoform | 2.88 | 0.96 | 1.53 | 0.97 |
| EUR | case-ctrl | adult | isoform | 1.06 | 1.05 | 1 | 0.99 |
| EUR | HC-BT1 | adult | isoform | 1.02 | 1.01 | 0.99 | 0.99 |
| EUR | HC-BT2 | adult | isoform | 1.04 | 1.03 | 1 | 1 |
| EUR | HC-BT3 | adult | isoform | 1.03 | 1 | 0.99 | 0.99 |
| EUR | BT1-BT2 | adult | isoform | 1.03 | 1.02 | 1 | 1 |
| EUR | BT1-BT3 | adult | isoform | 1.06 | 1.03 | 0.99 | 0.99 |
| EUR | BT2-BT3 | adult | isoform | 1.02 | 1.01 | 0.99 | 0.99 |
| AFR | case-ctrl | adult | isoform | 0.99 | 0.98 | 0.99 | 0.99 |
| AFR | HC-BT1 | adult | isoform | 1.02 | 0.99 | 1 | 0.99 |
| AFR | HC-BT2 | adult | isoform | 0.98 | 0.97 | 0.99 | 0.99 |
| AFR | HC-BT3 | adult | isoform | 0.97 | 0.96 | 0.99 | 0.99 |
| AFR | BT1-BT2 | adult | isoform | 0.98 | 0.96 | 0.98 | 0.98 |
| AFR | BT1-BT3 | adult | isoform | 0.99 | 0.99 | 0.99 | 1 |
| AFR | BT2-BT3 | adult | isoform | 0.99 | 0.98 | 0.99 | 0.99 |
| AMR | case-ctrl | adult | isoform | 1.32 | 0.98 | 1.11 | 0.99 |
| AMR | HC-BT1 | adult | isoform | 1.42 | 1.02 | 1.14 | 1 |
| AMR | HC-BT2 | adult | isoform | 1.21 | 0.99 | 1.05 | 1 |
| AMR | HC-BT3 | adult | isoform | 1.05 | 0.98 | 1 | 0.99 |
| AMR | BT1-BT2 | adult | isoform | 1.05 | 1.01 | 0.99 | 0.99 |
| AMR | BT1-BT3 | adult | isoform | 1.17 | 1.03 | 1.04 | 1 |
| AMR | BT2-BT3 | adult | isoform | 1.03 | 0.98 | 0.99 | 0.99 |

| | | | | | | | |
|---|---|---|---|---|---|---|---|
| EAS | case-ctrl | adult | isoform | 1.01 | 0.92 | 0.96 | 0.97 |
| EAS | HC-BT1 | adult | isoform | 0.99 | 0.92 | 0.94 | 0.96 |
| EAS | HC-BT2 | adult | isoform | 1.1 | 0.98 | 0.97 | 0.99 |
| EAS | HC-BT3 | adult | isoform | 0.99 | 0.96 | 0.95 | 0.98 |
| EAS | BT1-BT2 | adult | isoform | 1.07 | 0.91 | 0.93 | 0.98 |
| EAS | BT1-BT3 | adult | isoform | 1.05 | 0.89 | 0.94 | 0.96 |
| EAS | BT2-BT3 | adult | isoform | 1.08 | 0.92 | 0.94 | 0.97 |
| SAS | case-ctrl | adult | isoform | 1.03 | 0.98 | 0.96 | 0.99 |
| SAS | HC-BT1 | adult | isoform | 1.06 | 1.01 | 0.95 | 1 |
| SAS | HC-BT2 | adult | isoform | 1.02 | 1.01 | 0.9 | 1.01 |
| SAS | HC-BT3 | adult | isoform | 1.05 | 1 | 0.95 | 1 |
| SAS | BT1-BT2 | adult | isoform | 1.05 | 1.01 | 0.83 | 1.01 |
| SAS | BT1-BT3 | adult | isoform | 1.12 | 0.97 | 0.92 | 1.02 |
| SAS | BT2-BT3 | adult | isoform | 1.04 | 1.03 | 0.85 | 1.04 |

Legend: PCs = Principal Components. "noPCs" means association results without any PCs included in the logistic regression model. "withPCs" means association results with the first five genotype PCs included in the logistic regression model. The metric *Inflation* is calculated using a Bayesian method based on the empirical null distribution. The metric *Lambda* is the genomic inflation factor, which is calculated by comparing the observed distribution of p-values to the expected uniform distribution under the null hypothesis of no association. A value close to 1 for both metrics indicates that the distribution of p-values closely follows the expected uniform distribution, suggesting minimal inflation. This result shows there is an inflation when genotype PCs are not adjusted for isoform TWAS in adult brain in the combined multi-ancestry sample.

#### Table S11. TWAS of adult brain splicing expression: Inflation of results with and without genotype principal components (PCs) as covariates.

| Ancestry | Group | Brain Developmental Stage | Transcriptome | Lambda (noPCs) | Lambda (withPCs) | Inflation (noPCs) | Inflation (withPCs) |
|---|---|---|---|---|---|---|---|
| ALL | case-ctrl | adult | splicing | 2.04 | 1 | 1.36 | 0.97 |
| ALL | HC-BT1 | adult | splicing | 3.77 | 0.96 | 1.64 | 0.96 |
| ALL | HC-BT2 | adult | splicing | 2.36 | 0.96 | 1.42 | 0.96 |
| ALL | HC-BT3 | adult | splicing | 1.48 | 0.97 | 1.17 | 0.96 |
| ALL | BT1-BT2 | adult | splicing | 1.24 | 0.98 | 1.08 | 0.97 |
| ALL | BT1-BT3 | adult | splicing | 5.16 | 0.96 | 1.81 | 0.97 |
| ALL | BT2-BT3 | adult | splicing | 3.2 | 0.92 | 1.58 | 0.96 |
| EUR | case-ctrl | adult | splicing | 1.03 | 1.03 | 1 | 1 |
| EUR | HC-BT1 | adult | splicing | 1.02 | 0.99 | 0.99 | 0.99 |
| EUR | HC-BT2 | adult | splicing | 1.01 | 0.98 | 0.98 | 0.98 |
| EUR | HC-BT3 | adult | splicing | 1.08 | 1.08 | 1 | 1 |
| EUR | BT1-BT2 | adult | splicing | 0.98 | 0.97 | 0.97 | 0.97 |
| EUR | BT1-BT3 | adult | splicing | 1.05 | 1.01 | 1 | 0.99 |
| EUR | BT2-BT3 | adult | splicing | 0.99 | 0.97 | 0.97 | 0.97 |
| AFR | case-ctrl | adult | splicing | 1.01 | 1.01 | 0.98 | 0.98 |
| AFR | HC-BT1 | adult | splicing | 1.02 | 0.97 | 0.97 | 0.97 |
| AFR | HC-BT2 | adult | splicing | 1 | 1 | 0.98 | 0.99 |
| AFR | HC-BT3 | adult | splicing | 1.03 | 1.03 | 0.98 | 0.99 |
| AFR | BT1-BT2 | adult | splicing | 1.03 | 0.99 | 0.99 | 0.98 |
| AFR | BT1-BT3 | adult | splicing | 0.95 | 0.96 | 0.98 | 0.99 |
| AFR | BT2-BT3 | adult | splicing | 0.97 | 0.99 | 0.97 | 0.98 |
| AMR | case-ctrl | adult | splicing | 1.32 | 0.95 | 1.09 | 0.96 |
| AMR | HC-BT1 | adult | splicing | 1.44 | 1 | 1.14 | 0.98 |
| AMR | HC-BT2 | adult | splicing | 1.14 | 0.99 | 1.04 | 0.98 |
| AMR | HC-BT3 | adult | splicing | 1.03 | 0.97 | 0.99 | 0.98 |
| AMR | BT1-BT2 | adult | splicing | 1 | 0.96 | 0.98 | 0.99 |
| AMR | BT1-BT3 | adult | splicing | 1.11 | 0.96 | 1.03 | 0.98 |
| AMR | BT2-BT3 | adult | splicing | 1.03 | 0.98 | 0.98 | 0.98 |
| EAS | case-ctrl | adult | splicing | 1.02 | 0.98 | 0.96 | 0.97 |
| EAS | HC-BT1 | adult | splicing | 0.98 | 0.92 | 0.93 | 0.96 |
| EAS | HC-BT2 | adult | splicing | 1.09 | 0.99 | 0.96 | 0.97 |
| EAS | HC-BT3 | adult | splicing | 1.04 | 1.03 | 0.95 | 1 |
| EAS | BT1-BT2 | adult | splicing | 1.05 | 0.96 | 0.92 | 0.97 |
| EAS | BT1-BT3 | adult | splicing | 0.97 | 0.87 | 0.91 | 0.95 |
| EAS | BT2-BT3 | adult | splicing | 1.06 | 0.93 | 0.93 | 0.96 |
| SAS | case-ctrl | adult | splicing | 1.02 | 0.98 | 0.95 | 0.98 |

| | | | | | | | |
|---|---|---|---|---|---|---|---|
| SAS | HC-BT1 | adult | splicing | 1 | 0.94 | 0.93 | 0.98 |
| SAS | HC-BT2 | adult | splicing | 0.95 | 1.02 | 0.89 | 1 |
| SAS | HC-BT3 | adult | splicing | 1.04 | 1.02 | 0.94 | 1 |
| SAS | BT1-BT2 | adult | splicing | 0.95 | 0.99 | 0.81 | 1.01 |
| SAS | BT1-BT3 | adult | splicing | 1.11 | 1.07 | 0.91 | 1.05 |
| SAS | BT2-BT3 | adult | splicing | 0.99 | 1.1 | 0.84 | 1.07 |

Legend: PCs = Principal Components. "noPCs" means association results without any PCs included in the logistic regression model. "withPCs" means association results with the first five genotype PCs included in the logistic regression model. The metric *Inflation* is calculated using a Bayesian method based on the empirical null distribution. The metric *Lambda* is the genomic inflation factor, which is calculated by comparing the observed distribution of p-values to the expected uniform distribution under the null hypothesis of no association. A value close to 1 for both metrics indicates that the distribution of p-values closely follows the expected uniform distribution, suggesting minimal inflation. This result shows there is an inflation when genotype PCs are not adjusted for splicing TWAS in adult brain in the combined multi-ancestry sample.

#### Table S12. TWAS suggestive gene/transcript/splicing associations with the threshold FDR < 0.1.

| Original ID | Gene ID | Gene Name | Comparison | Developmental Stage | Transcriptome | Ancestry | Effect Size | SE | Z-score | P-value | FDR | N_samples |
|---|---|---|---|---|---|---|---|---|---|---|---|---|
| **ENSG00000165506** | **ENSG00000165506** | **DNAAF2** | **HC-BT2** | **adult** | **gene-level** | **SAS** | **5.72** | **0.89** | **6.46** | **3.32E-07** | **0.005** | **33** |
| **ENST00000608819** | **ENSG00000272941** | **novel transcript, antisense to C7orf49** | **HC-BT3** | **adult** | **isoform-level** | **AFR** | **-0.95** | **0.18** | **-5.28** | **2.40E-07** | **0.008** | **319** |
| **ENST00000481410** | **ENSG00000122783** | **CYREN** | **HC-BT3** | **adult** | **isoform-level** | **AFR** | **-0.89** | **0.18** | **-4.95** | **1.23E-06** | **0.016** | **319** |
| **ENST00000466307** | **ENSG00000146859** | **TMEM140** | **HC-BT3** | **adult** | **isoform-level** | **AFR** | **-1.08** | **0.22** | **-4.84** | **2.05E-06** | **0.016** | **319** |
| **ENST00000487774** | **ENSG00000122783** | **CYREN** | **HC-BT3** | **adult** | **isoform-level** | **AFR** | **-0.88** | **0.18** | **-4.83** | **2.12E-06** | **0.016** | **319** |
| **ENST00000275767** | **ENSG00000146859** | **TMEM140** | **HC-BT3** | **adult** | **isoform-level** | **AFR** | **-0.59** | **0.12** | **-4.82** | **2.28E-06** | **0.016** | **319** |
| **ENST00000670978** | **ENSG00000287733** | **novel transcript** | **HC-BT3** | **adult** | **isoform-level** | **AFR** | **0.29** | **0.06** | **4.71** | **3.64E-06** | **0.021** | **319** |
| **ENSG00000257176** | **ENSG00000257176** | **novel transcript, antisense to FAR2** | **HC-BT3** | **fetal** | **gene-level** | **AFR** | **0.74** | **0.16** | **4.75** | **3.08E-06** | **0.022** | **319** |
| **ENSG00000162817** | **ENSG00000162817** | **C1orf115** | **HC-BT1** | **adult** | **gene-level** | **SAS** | **-4.04** | **0.72** | **-5.63** | **2.38E-06** | **0.034** | **37** |
| **ENST00000414809** | **ENSG00000117407** | **ARTN** | **HC-BT2** | **adult** | **isoform-level** | **SAS** | **1.87** | **0.31** | **6.01** | **1.18E-06** | **0.040** | **33** |
| **ENSG00000287733** | **ENSG00000287733** | **novel transcript** | **HC-BT3** | **fetal** | **gene-level** | **AFR** | **1.03** | **0.23** | **4.45** | **1.19E-05** | **0.042** | **319** |
| **ENSG00000275476** | **ENSG00000275476** | **novel transcript, antisense to FAR2** | **HC-BT3** | **fetal** | **gene-level** | **AFR** | **2.13** | **0.49** | **4.33** | **1.97E-05** | **0.046** | **319** |
| ENSG00000228857 | ENSG00000228857 | ACTR3-AS1 | HC-BT3 | fetal | gene-level | EUR | 3.03 | 0.67 | 4.51 | 7.60E-06 | 0.053 | 669 |
| ENSG00000106483 | ENSG00000106483 | SFRP4 | BT1-BT3 | adult | gene-level | EUR | 2.28 | 0.49 | 4.68 | 3.78E-06 | 0.054 | 496 |
| ENSG00000143147 | ENSG00000143147 | GPR161 | BT1-BT2 | fetal | gene-level | SAS | 4.93 | 0.50 | 9.82 | 9.73E-06 | 0.068 | 10 |
| ENSG00000244063 | ENSG00000244063 | novel transcript | HC-BT3 | fetal | gene-level | EUR | 1.42 | 0.33 | 4.29 | 2.05E-05 | 0.072 | 669 |
| ENSG00000259954 | ENSG00000259954 | IL21R-AS1 | HC-BT1 | fetal | gene-level | AMR | -0.78 | 0.17 | -4.60 | 1.04E-05 | 0.073 | 122 |
| ENST00000483029 | ENSG00000122783 | CYREN | HC-BT3 | adult | isoform-level | AFR | 1.34 | 0.31 | 4.35 | 1.85E-05 | 0.090 | 319 |
| ENST00000210444 | ENSG00000095380 | NANS | HC-BT3 | adult | isoform-level | AFR | -2.79 | 0.65 | -4.31 | 2.17E-05 | 0.090 | 319 |
| ENST00000560055 | ENSG00000108107 | RPL28 | HC-BT3 | adult | isoform-level | AFR | -0.46 | 0.11 | -4.29 | 2.36E-05 | 0.090 | 319 |
| ENST00000674186 | ENSG00000151490 | PTPRO | HC-BT3 | adult | isoform-level | ALL | -0.33 | 0.07 | -4.71 | 2.76E-06 | 0.094 | 1200 |
| intron_7_151028610_151033443 | ENSG00000197150 | ABCB8 | HC-BT3 | adult | splicing-level | AFR | 1.10 | 0.25 | 4.43 | 1.31E-05 | 0.097 | 319 |
| intron_19_10295612_10296339 | ENSG00000105376 | ICAM5 | BT2-BT3 | adult | splicing-level | SAS | -2.64 | 0.34 | -7.87 | 1.36E-05 | 0.099 | 12 |

Legend: The gene/transcript/splicing associations with bold font are with the threshold Benjamini & Hochberg (BH) FDR < 0.05, which were also shown in the Main text Table 1. The 14 unique genes highlighted with blue color are detected as suggestively causal genes of Biotypes or psychosis disorders using MR-JTI based on Bonferroni correction of 19 total unique genes listed here. The “Original ID” were converted to “Gene

ID” and “Gene Name” through “g:Convert Gene ID conversion” function at g:Profiler website (https://biit.cs.ut.ee/gprofiler/convert)[1]. HC = Healthy Control, BT1 = Biotype 1, BT2 = Biotype 2, BT3 = Biotype 3. EUR = European, AFR = African, AMR = Admixed American, EAS = East Asian, SAS = South Asian.

**Table S13. Enriched biological pathways of the 14 unique causal genes based on suggestive TWAS associations in Table S12.**

| Pathway identifier | Pathway name | #Entities found | #Entities total | Entities ratio | Entities pValue | Entities FDR | #Reactions found | #Reactions total | Reactions ratio | Submitted entities found | Mapped entities |
|---|---|---|---|---|---|---|---|---|---|---|---|
| R-HSA-156902 | Peptide chain elongation | 2 | 90 | 0.008 | 0.006 | 0.029 | 4 | 5 | 3.38E-04 | ENSG00000108107 | P46776; P46779 |
| R-HSA-72764 | Eukaryotic Translation Termination | 2 | 94 | 0.008 | 0.006 | 0.029 | 3 | 5 | 3.38E-04 | ENSG00000108107 | P46776; P46779 |
| R-HSA-2408557 | Selenocysteine synthesis | 2 | 94 | 0.008 | 0.006 | 0.029 | 2 | 7 | 4.73E-04 | ENSG00000108107 | P46776; P46779 |
| R-HSA-156842 | Eukaryotic Translation Elongation | 2 | 95 | 0.008 | 0.006 | 0.029 | 4 | 9 | 6.09E-04 | ENSG00000108107 | P46776; P46779 |
| R-HSA-975956 | Nonsense Mediated Decay (NMD) independent of the Exon Junction Complex (EJC) | 2 | 96 | 0.008 | 0.006 | 0.029 | 1 | 1 | 6.76E-05 | ENSG00000108107 | P46776; P46779 |
| R-HSA-192823 | Viral mRNA Translation | 2 | 101 | 0.009 | 0.007 | 0.029 | 2 | 2 | 1.35E-04 | ENSG00000108107 | P46776; P46779 |
| R-HSA-72689 | Formation of a pool of free 40S subunits | 2 | 102 | 0.009 | 0.007 | 0.029 | 1 | 2 | 1.35E-04 | ENSG00000108107 | P46776; P46779 |
| R-HSA-9633012 | Response of EIF2AK4 (GCN2) to amino acid deficiency | 2 | 102 | 0.009 | 0.007 | 0.029 | 4 | 16 | 0.001 | ENSG00000108107 | P46776; P46779 |
| R-HSA-156827 | L13a-mediated translational silencing of Ceruloplasmin expression | 2 | 112 | 0.009 | 0.009 | 0.029 | 1 | 3 | 2.03E-04 | ENSG00000108107 | P46776; P46779 |
| R-HSA-1799339 | SRP-dependent cotranslational protein targeting to membrane | 2 | 113 | 0.010 | 0.009 | 0.029 | 5 | 5 | 3.38E-04 | ENSG00000108107 | P46776; P46779 |
| R-HSA-72706 | GTP hydrolysis and joining of the 60S ribosomal subunit | 2 | 113 | 0.010 | 0.009 | 0.029 | 2 | 3 | 2.03E-04 | ENSG00000108107 | P46776; P46779 |
| R-HSA-927802 | Nonsense-Mediated Decay (NMD) | 2 | 117 | 0.010 | 0.009 | 0.029 | 5 | 6 | 4.06E-04 | ENSG00000108107 | P46776; P46779 |
| R-HSA-975957 | Nonsense Mediated Decay (NMD) enhanced by the Exon Junction Complex (EJC) | 2 | 117 | 0.010 | 0.009 | 0.029 | 4 | 5 | 3.38E-04 | ENSG00000108107 | P46776; P46779 |
| R-HSA-2408522 | Selenoamino acid metabolism | 2 | 118 | 0.010 | 0.010 | 0.029 | 2 | 23 | 0.002 | ENSG00000108107 | P46776; P46779 |
| R-HSA-72613 | Eukaryotic Translation Initiation | 2 | 120 | 0.010 | 0.010 | 0.030 | 4 | 21 | 0.001 | ENSG00000108107 | P46776; P46779 |

| | | | | | | | | | | | |
|---|---|---|---|---|---|---|---|---|---|---|---|
| R-HSA-72737 | Cap-dependent Translation Initiation | 2 | 120 | 0.010 | 0.010 | 0.030 | 3 | 18 | 0.001 | ENSG00000108107 | P46776; P46779 |
| R-HSA-168273 | Influenza Viral RNA Transcription and Replication | 2 | 152 | 0.013 | 0.015 | 0.032 | 2 | 13 | 8.79E-04 | ENSG00000108107 | P46776; P46779 |
| R-HSA-9711097 | Cellular response to starvation | 2 | 157 | 0.013 | 0.016 | 0.032 | 4 | 28 | 0.002 | ENSG00000108107 | P46776; P46779 |
| R-HSA-9010553 | Regulation of expression of SLITs and ROBOs | 2 | 159 | 0.013 | 0.017 | 0.032 | 2 | 19 | 0.001 | ENSG00000108107 | P46776; P46779 |
| R-HSA-168255 | Influenza Infection | 2 | 172 | 0.015 | 0.020 | 0.032 | 2 | 58 | 0.004 | ENSG00000108107 | P46776; P46779 |
| R-HSA-6791226 | Major pathway of rRNA processing in the nucleolus and cytosol | 2 | 183 | 0.015 | 0.022 | 0.032 | 1 | 7 | 4.73E-04 | ENSG00000108107 | P46776; P46779 |
| R-HSA-9034015 | Signaling by NTRK3 (TRKC) | 1 | 19 | 0.002 | 0.024 | 0.032 | 1 | 19 | 0.001 | ENSG00000151490 | Q16827 |
| R-HSA-8868773 | rRNA processing in the nucleus and cytosol | 2 | 193 | 0.016 | 0.024 | 0.032 | 1 | 15 | 0.001 | ENSG00000108107 | P46776; P46779 |
| R-HSA-9629569 | Protein hydroxylation | 1 | 20 | 0.002 | 0.025 | 0.032 | 1 | 10 | 6.76E-04 | ENSG00000108107 | P46776 |
| R-HSA-72312 | rRNA processing | 2 | 203 | 0.017 | 0.027 | 0.032 | 1 | 21 | 0.001 | ENSG00000108107 | P46776; P46779 |
| R-HSA-376176 | Signaling by ROBO receptors | 2 | 205 | 0.017 | 0.027 | 0.032 | 2 | 59 | 0.004 | ENSG00000108107 | P46776; P46779 |
| R-HSA-422475 | Axon guidance | 3 | 540 | 0.046 | 0.029 | 0.032 | 22 | 294 | 0.020 | ENSG00000108107; ENSG00000117407 | P46776; Q5T4W7; P46779 |
| R-HSA-9675108 | Nervous system development | 3 | 566 | 0.048 | 0.032 | 0.032 | 22 | 320 | 0.022 | ENSG00000108107; ENSG00000117407 | P46776; Q5T4W7; P46779 |
| **R-HSA-8853659** | **RET signaling** | **1** | **41** | **0.003** | **0.051** | **0.051** | **19** | **24** | **0.002** | **ENSG00000117407** | **Q5T4W7** |
| **R-HSA-419037** | **NCAM1 interactions** | **1** | **42** | **0.004** | **0.052** | **0.052** | **1** | **10** | **6.76E-04** | **ENSG00000117407** | **Q5T4W7** |
| R-HSA-72766 | Translation | 2 | 294 | 0.025 | 0.052 | 0.052 | 16 | 99 | 0.007 | ENSG00000108107 | P46776; P46779 |
| R-HSA-71291 | Metabolism of amino acids and derivatives | 2 | 355 | 0.030 | 0.073 | 0.073 | 2 | 249 | 0.017 | ENSG00000108107 | P46776; P46779 |
| R-HSA-163841 | Gamma carboxylation, hypusinylation, hydroxylation, and arylsulfatase activation | 1 | 61 | 0.005 | 0.075 | 0.075 | 1 | 50 | 0.003 | ENSG00000108107 | P46776 |
| **R-HSA-375165** | **NCAM signaling for neurite out-growth** | **1** | **64** | **0.005** | **0.078** | **0.078** | **1** | **23** | **0.002** | **ENSG00000117407** | **Q5T4W7** |

Legend: The significance threshold for the biological pathway enrichment analysis is Benjamini & Hochberg (BH) FDR < 0.1. The biological pathway enrichment analysis was implemented by Reactome (https://reactome.org/)[2]. The three pathways with bold font were also significant (P < 0.05) for the seven putatively causal genes/transcripts, which were shown in the Main text Figure 7.

### Table S14. Resampling analysis of TWAS findings.

| Gene Id | Gene Name | Effect Size | SE | Z-score | P-value | N_samples | FDR | Developmental Stage | Transcriptome | Comparison | Ancestry | Resampling Dataset |
|---|---|---|---|---|---|---|---|---|---|---|---|---|
| ENSG00000162817 | C1orf115 | -4.036 | 0.717 | -5.629 | 2.38e-06 | 37 | 0.034 | adult | gene-level | HC-BT1 | SAS | original |
| | | -3.911 | 0.707 | -5.529 | 6.54e-06 | 30 | 0.031 | | | | | r4 |
| ENSG00000165506 | DNAAF2 | 5.719 | 0.885 | 6.462 | 3.32e-07 | 33 | 0.005 | adult | gene-level | HC-BT2 | SAS | original |
| | | 7.787 | 0.911 | 8.547 | 2.81e-08 | 23 | 4.39e-05 | | | | | r6 |
| | | 7.493 | 0.801 | 9.349 | 4.16e-10 | 30 | 5.87e-06 | | | | | r8 |
| | | 7.643 | 0.865 | 8.837 | 7.47e-09 | 25 | 1.05e-04 | | | | | r9 |
| | | 8.041 | 0.797 | 10.092 | 6.43e-10 | 25 | 9.06e-06 | | | | | r10 |
| ENSG00000257176 | novel transcript, antisense to FAR2 | 0.739 | 0.156 | 4.750 | 3.08e-06 | 319 | 0.022 | fetal | gene-level | HC-BT3 | AFR | original |
| | | 0.890 | 0.166 | 5.360 | 1.89e-07 | 252 | 0.001 | | | | | r1 |
| | | 0.768 | 0.167 | 4.588 | 7.00e-06 | 258 | 0.035 | | | | | r3 |
| | | 0.803 | 0.175 | 4.596 | 6.82e-06 | 253 | 0.048 | | | | | r8 |
| | | 0.803 | 0.171 | 4.682 | 4.64e-06 | 254 | 0.033 | | | | | r10 |
| ENSG00000275476 | novel transcript, antisense to FAR2 | 2.128 | 0.491 | 4.334 | 1.97e-05 | 319 | 0.046 | fetal | gene-level | HC-BT3 | AFR | original |
| | | 2.831 | 0.529 | 5.357 | 1.92e-07 | 252 | 0.001 | | | | | r1 |
| | | 2.386 | 0.529 | 4.507 | 1.00e-05 | 258 | 0.035 | | | | | r3 |
| ENST00000275767 | TMEM140 | -0.587 | 0.122 | -4.816 | 2.28e-06 | 319 | 0.016 | adult | isoform-level | HC-BT3 | AFR | original |
| | | -0.618 | 0.135 | -4.571 | 7.59e-06 | 256 | 0.039 | | | | | r2 |
| | | -0.637 | 0.136 | -4.702 | 4.24e-06 | 254 | 0.036 | | | | | r6 |
| | | -0.660 | 0.135 | -4.884 | 1.85e-06 | 253 | 0.014 | | | | | r8 |
| ENST00000414809 | ARTN | 1.872 | 0.311 | 6.012 | 1.18e-06 | 33 | 0.040 | adult | isoform-level | HC-BT2 | SAS | original |
| | | 1.948 | 0.213 | 9.126 | 6.21e-09 | 24 | 2.12e-04 | | | | | r5 |
| | | 1.971 | 0.200 | 9.843 | 1.03e-09 | 25 | 3.52e-05 | | | | | r9 |

| | | | | | | | | | | | | |
|---|---|---|---|---|---|---|---|---|---|---|---|---|
| | | 2.018 | 0.192 | 10.499 | 3.03e-10 | 25 | 1.03e-05 | | | | | r10 |
| ENST00000466307 | TMEM140 | -1.084 | 0.224 | -4.838 | 2.05e-06 | 319 | 0.016 | adult | isoform-level | HC-BT3 | AFR | original |
| | | -1.136 | 0.248 | -4.577 | 7.39e-06 | 256 | 0.039 | | | | | r2 |
| | | -1.175 | 0.249 | -4.727 | 3.80e-06 | 254 | 0.036 | | | | | r6 |
| | | -1.214 | 0.248 | -4.890 | 1.80e-06 | 253 | 0.014 | | | | | r8 |
| ENST00000481410 | CYREN | -0.885 | 0.179 | -4.945 | 1.23e-06 | 319 | 0.016 | adult | isoform-level | HC-BT3 | AFR | original |
| | | -0.917 | 0.201 | -4.568 | 7.70e-06 | 256 | 0.039 | | | | | r2 |
| | | -0.927 | 0.200 | -4.645 | 5.47e-06 | 254 | 0.037 | | | | | r6 |
| | | -0.969 | 0.198 | -4.902 | 1.70e-06 | 253 | 0.014 | | | | | r8 |
| ENST00000487774 | CYREN | -0.880 | 0.182 | -4.831 | 2.12e-06 | 319 | 0.016 | adult | isoform-level | HC-BT3 | AFR | original |
| | | -0.925 | 0.203 | -4.561 | 7.94e-06 | 256 | 0.039 | | | | | r2 |
| | | -0.951 | 0.202 | -4.710 | 4.10e-06 | 254 | 0.036 | | | | | r6 |
| | | -0.980 | 0.202 | -4.859 | 2.08e-06 | 253 | 0.014 | | | | | r8 |
| ENST00000608819 | novel transcript, antisense to C7orf49 | -0.953 | 0.180 | -5.280 | 2.40e-07 | 319 | 0.008 | adult | isoform-level | HC-BT3 | AFR | original |
| | | -0.927 | 0.198 | -4.680 | 4.71e-06 | 252 | 0.032 | | | | | r1 |
| | | -0.999 | 0.202 | -4.956 | 1.31e-06 | 256 | 0.015 | | | | | r2 |
| | | -0.985 | 0.201 | -4.905 | 1.67e-06 | 254 | 0.036 | | | | | r6 |
| | | -1.035 | 0.199 | -5.198 | 4.18e-07 | 253 | 0.014 | | | | | r8 |
| ENST00000670978 | novel transcript | 0.285 | 0.061 | 4.714 | 3.64e-06 | 319 | 0.021 | adult | isoform-level | HC-BT3 | AFR | original |
| | | 0.315 | 0.067 | 4.700 | 4.28e-06 | 253 | 0.024 | | | | | r8 |

Legend: Resampling was used to test the robustness of the findings from TWAS analyses of psychosis Biotypes. We randomly selected 80% of the whole sample for 10 times, and each time we performed all the TWAS analyses. We then evaluated the robustness of our findings by comparing the results from resampling and from our main analyses. “original” means the initial test statistics from our main TWAS analyses. “r1” … “r10” represent the iterations from 1 to 10 in the resampling process. Only significant (BH FDR < 0.05) associations were shown here. Resampling showed the TWAS results of eleven genes and transcripts (except for ENSG00000287733) replicated 1 to 4 times.

### Supplementary Figures

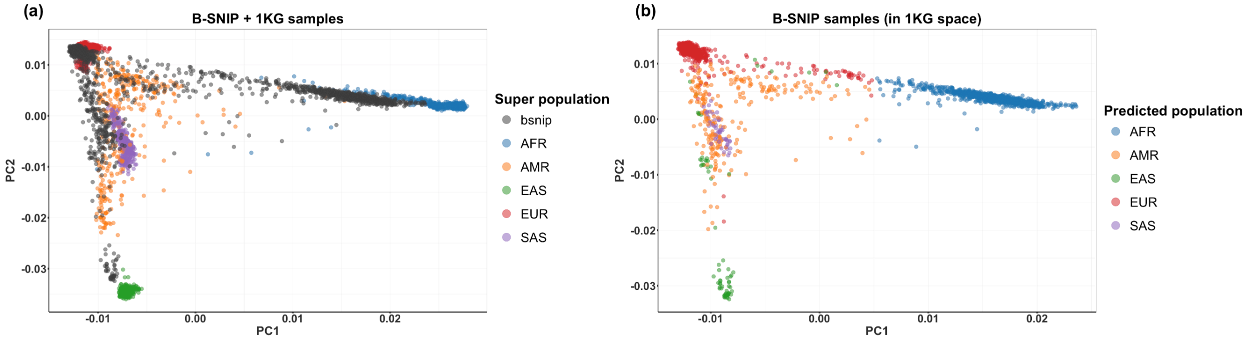


**Figure S1. B-SNIP individuals (N = 2,505) projected in 1KG Principal Components (PCs) 1 and 2 graphic space.** Legend: (a) shows a plot of the first 2 genotype PCs of the merged samples (B-SNIP + 1KG). (b) shows a plot of the first 2 genotype PCs of the target samples (B-SNIP) in the reference (1KG) PC space. The first 10 PCs were used when assigning ancestries using Random Forest (RF) method. There are five ancestries assigned based on the RF prediction method: EUR (N = 1234), AFR (N = 908), AMR (N = 237), EAS (N = 73) and SAS (N = 53). EUR = European, AFR = African, AMR = Admixed American, EAS = East Asian, SAS = South Asian.

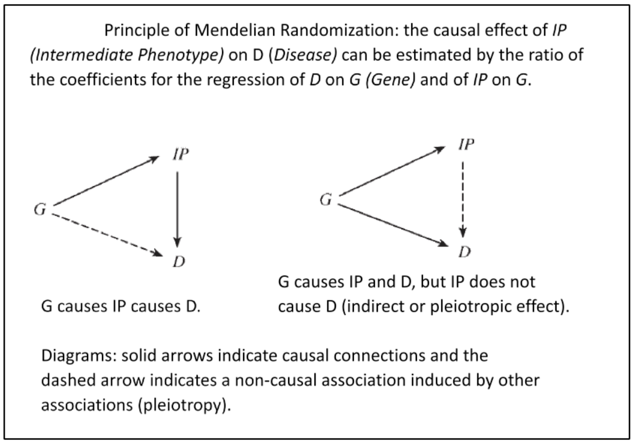


**Figure S2. Principle of Mendelian Randomization (from Thomas and Conti[3]).**

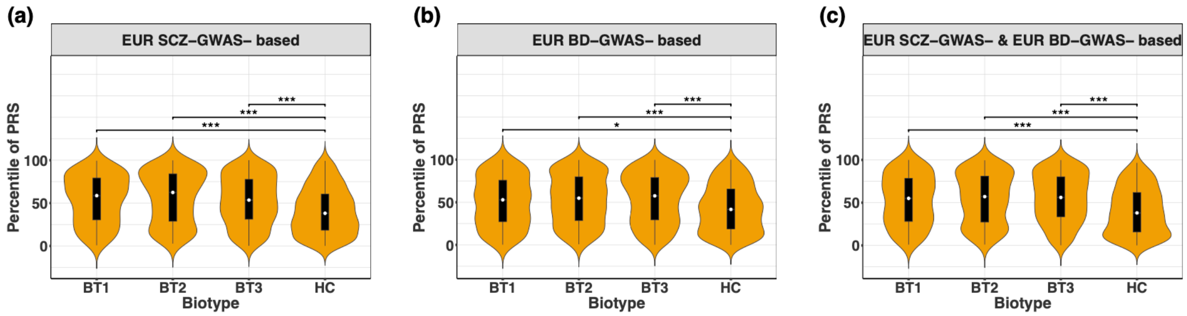


**Figure S3. Biotype vs. healthy control and Biotype vs. Biotype differences in AAPRSs based on SCZ-, BD- or both SCZ- and BD- GWAS summary statistics within EUR ancestry B-SNIP participants.** AAPRSs refer to Ancestry-Adjusted Polygenic Risk Score with *post hoc* ancestry adjustment of Khera. Each possible pair of Biotypes was tested for differences in AAPRSs, and each Biotype (BT) was compared with Healthy Control (HC). (a) Violin plot of EUR SCZ GWAS-summary statistics-based AAPRSs among Biotypes and with Healthy Control in EUR samples in the Combined Diagnosis group. (b) (c) Same comparisons of EUR BD GWAS-summary statistics-based AAPRSs, and EUR SCZ GWAS-summary statistics- & EUR BD GWAS-summary statistics- based AAPRSs. EUR = European ancestry. Wilcoxon tests were used for comparisons. Bonferroni-corrected significance threshold over 6 two-sample Wilcoxon tests for each ancestry is P-value < 8.33e-03. Only significant comparison results are labeled with asterisks. *** indicates P-value < 1.67e-04; ** indicates P-value < 1.67e-03; * indicates P-value < 8.33e-03. PGC 3 EUR SCZ GWAS summary statistics and PGC EUR BD GWAS summary statistics were used for PRS construction.

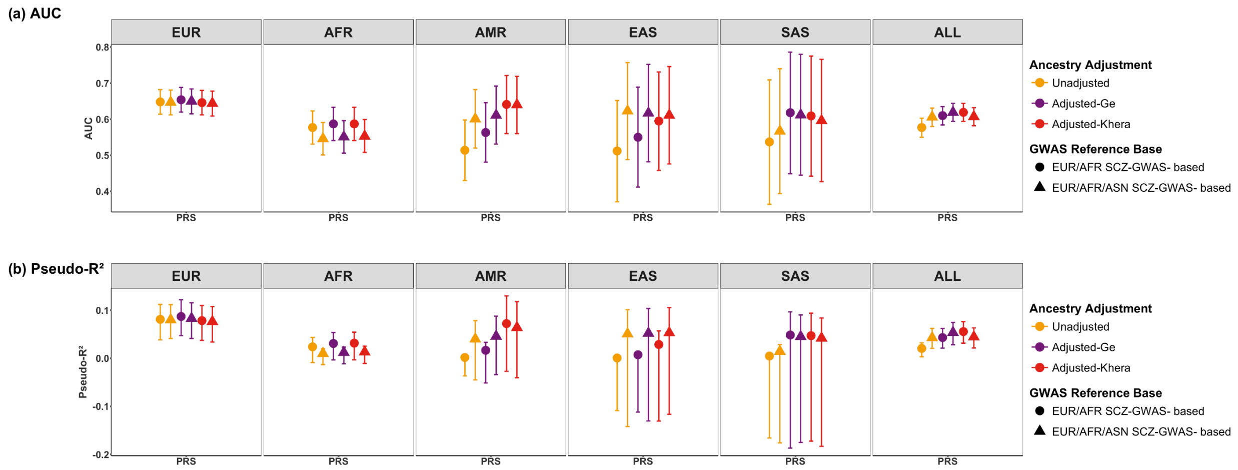


**Figure S4. Prediction accuracy of PRSs for case-control status before and after ancestry adjustment within and across 5 ancestries based on different GWAS summary statistics.** (a) The area under the receiver operating characteristic (ROC) curve (AUC) of PRSs. (b) The proportion of the case-control variance (Nagelkerke's pseudo-R2) explained by PRSs. Lines for Nagelkerke's pseudo-R2 in (b) correspond to 95% confidence intervals calculated via 1000 bootstrapping. The five ancestries were assigned by Random Forest inferred method based on 1KG reference. EUR = European, AFR = African, AMR = Admixed American, EAS = East Asian, SAS = South Asian, ALL = Combined multi-ancestry individuals of all the five ancestries. "Unadjusted" risk scores are the --meta option results from PRS-CSx prior to post hoc ancestry adjustment, and "Adjusted" refers to AAPRS (Ancestry-Adjusted Polygenic Risk Score) with post hoc ancestry adjustment of Khera or Ge. We find no overall advantage for either adjustment method or with the inclusion of Asian SCZ GWAS summary statistics in the prediction performance of case-control status.
